# High diversity amongst African *Treponema pallidum* genomes provides a window into global transmission dynamics of syphilis: A genomic epidemiology study

**DOI:** 10.64898/2026.03.20.26348644

**Authors:** Mathew A Beale, Michael Marks, Sarah Burl, Kirsty E Ambridge, Ethel Dauya, Aamirah Mussa, Bame Bame, Sikhulile Moyo, Michael Owusu, David P Kateete, Rogers Kamulegeya, Edgar Kigozi, Denis Kimbugwe, Becca L Handley, Mahlape P Mahlangu, Johanna ME Venter, Bianca Da Costa Dias, Yaw Adu-Sarkodie, Chelsea Morroni, Edith Nakku-Joloba, Rashida A Ferrand, Etienne E Müller, Nicholas R Thomson

## Abstract

**Background:** Global syphilis rates have risen dramatically since the early 2000s. Genomes can be used to inform rational control and intervention strategies by enhancing surveillance and ensuring vaccines have broad global utility. However, although >1000 *Treponema pallidum* genomes are now available from high-income countries, genomic data from Africa remain limited.

**Methods:** We combined samples from 1198 participants recruited into a genital ulcer aetiology study in Botswana, Ghana, Uganda and Zimbabwe (collected 2022-2023) with 276 samples from national syphilis surveillance in South Africa to generate 147 novel African *T. pallidum* genomes (collected 2006-2023). Combining these with 167 publicly available African genomes and 1062 genomes from 24 non-African countries, we performed contextual population genomic analyses to understand the *T. pallidum* genomic diversity and transmission within and between African countries and the rest of the world.

**Findings:** Contrasting with previous studies showing global circulation of highly similar *T. pallidum*, we found remarkable diversity amongst African *T. pallidum*. Of 56 sublineages, 20 were exclusively found amongst 6 African countries, 31 were found amongst 24 non-African countries, and 5 were found in both. Sublineage sharing between Africa and the rest of the world was rare, with 83.8% of African syphilis caused by locally circulating sublineages. Only 20.1% of African syphilis was resistant to macrolides (global average = 68.6%); where resistance occurred, this was strongly linked to introduction of global sublineages into Africa.

**Interpretation:** African *T. pallidum* is characterised by locally circulating strains not found globally. Since sublineage sharing between countries is low, cataloguing African *T. pallidum* diversity will require intense local sampling in many countries. These findings will inform ongoing strategies for genomic surveillance and vaccine design, whilst contributing to our understanding of the spread of antimicrobial resistance in Africa, enabling refined treatment guidelines based on local data.

**Funding:** Wellcome and the Gates Foundation.

## Introduction

Syphilis, caused by *Treponema pallidum* subspecies *pallidum* (TPA), is a major sexually transmitted infection (STI) causing 8 million new cases each year globally, and over 200,000 congenital syphilis related deaths^1–3^. Cases have been steadily rising in high-income countries (HICs) since the early 2000s, with some countries now experiencing their highest syphilis rates since the Second World War^4^.

TPA genomes can be divided into two deep-branching phylogenetic lineages (Nichols and SS14)^5^ which in turn can be subdivided into fine-scale sublineages. These sublineages provide sufficient granularity to partially resolve regional, national and global patterns of spread, and determine the contribution of any new lineage to overall genetic diversity^5,6^. However, the population structure of HICs has shown remarkable genomic homogeneity, with closely related or identical genomes found to have globally disseminated relatively recently^5,7,8^. Given the vast majority of TPA genomes sequenced to date have been collected in HICs^5^, there are substantial data gaps for Lower- and Middle-Income Countries (LMICs). These gaps are especially problematic in Africa, where syphilis rates are the highest in the world.

Although first-line treatment benzathine penicillin & (BPG) remains highly effective, there is a limited repertoire of alternative antimicrobials available for syphilis. Molecular and genomic surveillance data demonstrates most syphilis circulating in HICs is now resistant to macrolides^5,7,9^. Whether similar resistance is seen in LMICs and the extent to which this is influenced by introduction from HICs remains unknown but is critical to informing local and WHO guidelines. Syphilis genomics is also being used to inform vaccine design^10^. As the highest burden of sexually transmitted and congenital disease occurs in low-income settings^1,2,11^, it is critical to catalogue the full repertoire of antigenic diversity^6,10,12^, which will remain impossible without adequate data from LMIC settings.

We hypothesised that bacterial diversity, antimicrobial resistance patterns and transmission dynamics observed globally would differ in Africa and aimed to address this through population genomics of African TPA genomes. We established clinical pathways for molecular surveillance of genital ulcer disease in four African countries and performed whole genome sequencing of TPA PCR-positive samples to analyse the phylogeographical relationships between African and globally circulating TPA.

## Methods

### Samples

From July 2022 to December 2023, individuals presenting with genital ulcer(s) were recruited from primary care clinics in Botswana (eleven Gaborone District Health Management Team clinics), Ghana (four clinics in Kumasi, Sekondi, Accra, Tamale), Uganda (one Kampala clinic), and Zimbabwe (three Harare clinics) as part of the Multi-Country Aetiology of Genital Ulcer Study (MAGUS). In Botswana, participants were also reached through a social media advert, whilst radio spots were used in Ghana. From each participant, we collected detailed socio-demographic and clinical data including HIV status and previous history of genital ulcer disease. A multiplex PCR for HSV, *T. pallidum* and *Haemophilus ducreyi* was used to test all cases.

South African samples were collected as part of microbiological sentinel STI surveillance conducted between 2006 and 2023 from eight different provinces in South Africa. Samples were not consistently collected for individual regions, except for in Gauteng, where a routine surveillance programme was established at a single clinic in Alexandra, Johannesburg from 2007-2023, enabling consistent and systematic sampling for this province. Similar programmes were established in Western Cape in 2019 and KwaZulu-Natal in 2020. A search of samples from these programmes identified 276 *T. pallidum* PCR positive samples collected between 2006-2023.

### Ethics and Governance

Ethical approval for the MAGUS study was granted by the London School of Hygiene and Tropical Medicine Observational Research Ethics Committee (26731), the Health Research Development Committee of the Botswana Ministry of Health (HRDC#00975), the Committee on Human Research, Publication and Ethics, Kwame Nkrumah University of Science and Technology, Kumasi (CHRPE/AP/331/22), the School of Biomedical Sciences REC, Makerere University (SBS-2022-217), the Medical Research Council of Zimbabwe, the Biomedical Research and Training Institute IRB and Harare City Health Services (MRCZ/A/2878). Ethical approval for samples from South Africa was granted by the University of the Witwatersrand Human Research Ethics Committee (Medical) (M230157).

Samples were shipped to the UK for sequencing under Nagoya-compatible Access Benefit Sharing agreements.

### Sequencing, Bioinformatics and Phylogenetics

For archived South Africa, repeat testing of qPCR was performed and those with qPCR Cq <33 were submitted for whole genome sequencing. The prospectively recruited African samples were sequenced as available regardless of qPCR Cq. Samples were sequenced at the Wellcome Sanger Institute on Illumina NovaSeq 6000 after pooled sequence capture (Agilent SureSelect XT) and analysed using previously described bioinformatic pipelines^7,8^ (see Supplementary Methods).

## Results

### Contextualising African *T. pallidum* diversity

We prospectively recruited 1122 patients with genital ulcers from 2022-2023 in four African countries (Botswana, Ghana, Uganda, Zimbabwe) of which 80 (7.1%) genital ulcer swabs were positive for *T. pallidum*. Results of aetiological testing will be reported elsewhere, but briefly we found 75 (15. 4%) *T. pallidum* positive samples in Zimbabwe, 2 (1.1%) in Botswana, 1 (0.5%) in Ghana and 2 (0.8%) in Uganda. We further included a longitudinal collection of 276 *T. pallidum* positive swab DNAs collected between 2006-2023 by the National Institute for Communicable Diseases (NICD) in South Africa. We successfully recovered 147 genomes (meeting our minimum quality threshold of ≥75% of reference sites with ≥5X coverage) across the five countries. This includes the first TPA genomes to our knowledge from Botswana (n=2) and Uganda (n=1), whilst increasing the number of genomes from South Africa from 1 to 102, and in Zimbabwe from 12 to 55. No genomes were recovered from Ghana. The new African genomes were combined with previously published genomes from Africa (n=167, comprising 80 from Madagascar, 74 from Malawi, 12 from Zimbabwe and 1 from South Africa). In total, the earliest available African genomes included originated from Madagascar (2000-2007); the deepest longitudinal sampling was represented by South African genomes (2006-2023, including the only African genomes currently available from between 2008-2014); with more restricted sampling in Zimbabwe (2015, 2022-2023), Malawi (2020-2021), Botswana (2023) and Uganda (2023).

For context we included a set of genomes that represent the known global diversity of TPA from: Asia (n=151), the Caribbean (n=4), Europe (n=318), North America (n=201), Oceania (n=366), and South America (n=22)^5,6,10,14^. The final dataset, comprising 1376 genomes (Supplementary Figure 1), was used to infer a whole genome recombination-masked maximum likelihood phylogeny (see Supplementary Methods). All genomes were phylogenetically classified into major TPA lineages (Nichols or SS14), with 25.1% (345/1376) belonging to the Nichols-lineage and 74.9% (1031/1376) to SS14-lineage (Supplementary Figure 1, Supplementary Figure 2). In Africa, 40.8% (128/314) of genomes were Nichols-lineage, with 59.2% (186/314) of genomes in the SS14-lineage. Although Nichols-lineage was more common in Africa than elsewhere, this was strongly influenced by 80 genomes from Madagascar collected between 2000 and 2007, of which 98.8% (79/80) were Nichols-lineage (Supplementary Figure 3). In contrast, samples from other African countries such as Malawi (14.9% Nichols, 11/74), South Africa (27.5% Nichols, 28/102), and Zimbabwe (16.4% Nichols, 9/55) collected between 2006 and 2023 had SS14- and Nichols-lineage proportions similar to those globally^5^ and consistent with a previous African typing study^15^. We also had two genomes from Botswana (both SS14-lineage) and one from Uganda (Nichols-lineage).

To examine the historical spread of syphilis in Africa we defined fine-scale sublineages using rPinecone^16^, clustering genomes within 10 SNPs distance from a common ancestral node, as described previously^5^. Using this uniform approach to defining genetic sublineages, we defined 56 sublineages (comprising 2-519 genomes each) and 61 singletons. Of these, 26 sublineages fell within Nichols-lineage and 30 sublineages within SS14-lineage. We performed ancestral state reconstruction on our whole genome phylogeny, inferring the most recent common ancestors (MRCA) of African genomes in our maximum likelihood phylogeny (Figure 1A). Twenty-two sublineages had an African MRCA and belonged to four major African clades within SS14-lineage and three within Nichols-lineage (Figure 1B, Supplementary Figure 4) which were labelled according to the ancestral node (a1497, a1584, a1640, a1746, a2113, a2216, a2750). Clade a1746 comprised 6/25 African sublineages (4, 13, 23, 24, 27, 28) descended from a single SS14-lineage MRCA in Africa, representing 39.2% (123/314) of African genomes in the full dataset of 1376 genomes (Supplementary Figure 5). Two of the Nichols-lineage clades (a1584, a1640) comprised four (41, 42, 45, 49, 55) and six (40, 43, 46, 50, 52, 53, 60, 61) sublineages and represented 18.2% (57/314) and 21.7% (68/314) of African genomes respectively. The four remaining clades (a1497, a2113, a2216, a2750) each comprised one sublineage within either SS14- or Nichols-lineages (Figure 1B). Next, by subsampling a representative dataset of 440 SS14-lineage genomes (see methods) we inferred both the continental location and dates of ancestral nodes within SS14-lineage (Supplementary Figure 6; there was insufficient temporal signal to do this for Nichols-lineage - see Supplementary Figure 7). The largest African SS14-lineage clade had a median MRCA date of 1988 (95% highest posterior distribution 1980-1996, Supplementary Figure 8A), with two other African clades having MRCAs dated in the 2000s (Supplementary Figure 8C-G).

**Figure 1.**
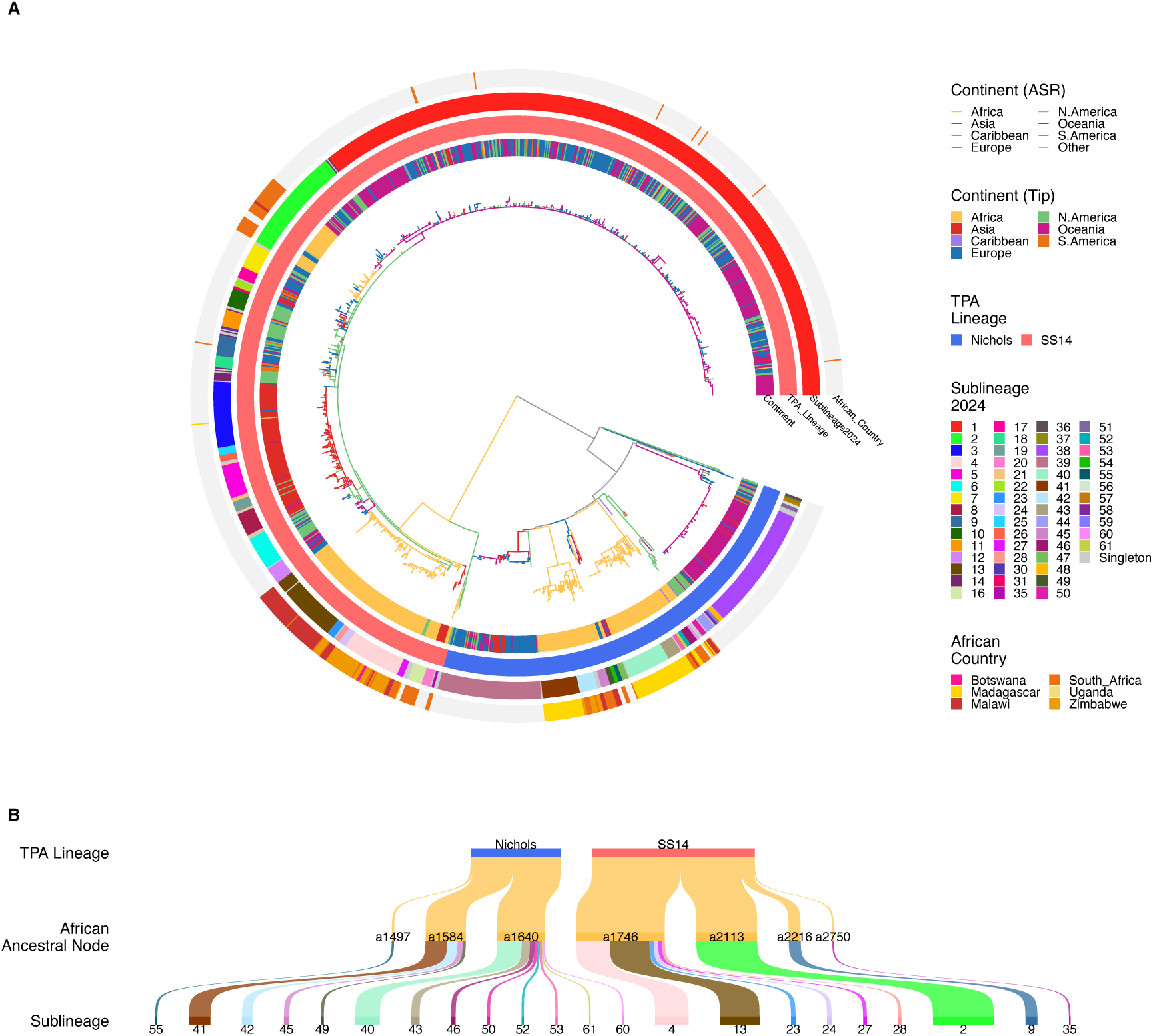
Phylogenetic analysis of *T. pallidum* lineage dispersal. A - Maximum likelihood whole genome phylogenies of 1376 T. pallidum subspecies pallidum genomes. Branches are coloured based on ancestral state reconstruction (ASR) of Continental location, and branch lengths are scaled by substitutions. Coloured tracks indicate Continent, Lineage, Sublineage, and African Country (where sample is from Africa). B – Sankey diagram showing sublineages descending from common ancestral nodes occurring in Africa.

The African clades could be divided into those containing sublineages that were globally distributed and those that were not. Unlike other settings where TPA sublineages are mostly globally distributed^5^, this was not the case for Africa – only 5/56 sublineages were found in both Africa and globally (representing 5/25 African sublineages, comprising 16.2% (51/314) of African samples). Of these, sublineages 1, 2 and 3 (all belonging to SS14-lineage) were also the most prevalent globally. We found that the globally dominant sublineage 1 was independently imported into South Africa at least four times (Supplementary Figure 9). Sublineage 2, present in North America, Europe and Australia, was also found as a monophyletic African clade in South Africa (38/82) and Malawi (2/82), which we predict was imported as a single introduction around 2005 (95% highest posterior distribution 2001-2008, Supplementary Figure 8C). Conversely, Sublineage 3 was largely restricted to Japan (n=49), with sporadic cases in Madagascar and the UK.

In South Africa, where we conducted longitudinal sampling from 2006-2023, the proportion of genomes belonging to globally shared sublineages increased from 5.0% (1/20) between 2007-2012 to 56.1% (46/82) from 2013-2023 (Supplementary Figure 6). Whilst we did not have detailed longitudinal sampling from other African countries, the proportion of samples from globally shared sublineages was just 1.9% (4/212), suggesting importation into these countries has been less pronounced. Consistent with this, over half (31/56) of sublineages detected globally were absent from our African dataset, with 20/56 sublineages thus far exclusive to Africa (Figure 2A, 2B) and nearly as many sublineages in the 6 African countries (25 sublineages) as in the rest of the world (24 countries, 36 sublineages) combined. Of note were the three African genomes (Sublineage 35) with an MRCA in the 1980s that were closely related to an SS14-lineage previously thought to be unique to Mexico^17^ (Mexico_A) and the USA^6^ (MD18Be and MD06B) (Supplementary Figure 8B, Supplementary Figure 10). To further understand the relevance of the Africa TPA we extrapolated the sublineage richness for Africa and the rest of the world. Our accumulation curves showed that whilst discovery of new TPA sublineages is starting to plateau globally, this is not the case for Africa where there is a significantly higher discovery rate of sublineages and singletons (Figure 2C, 2D).

**Figure 2.**
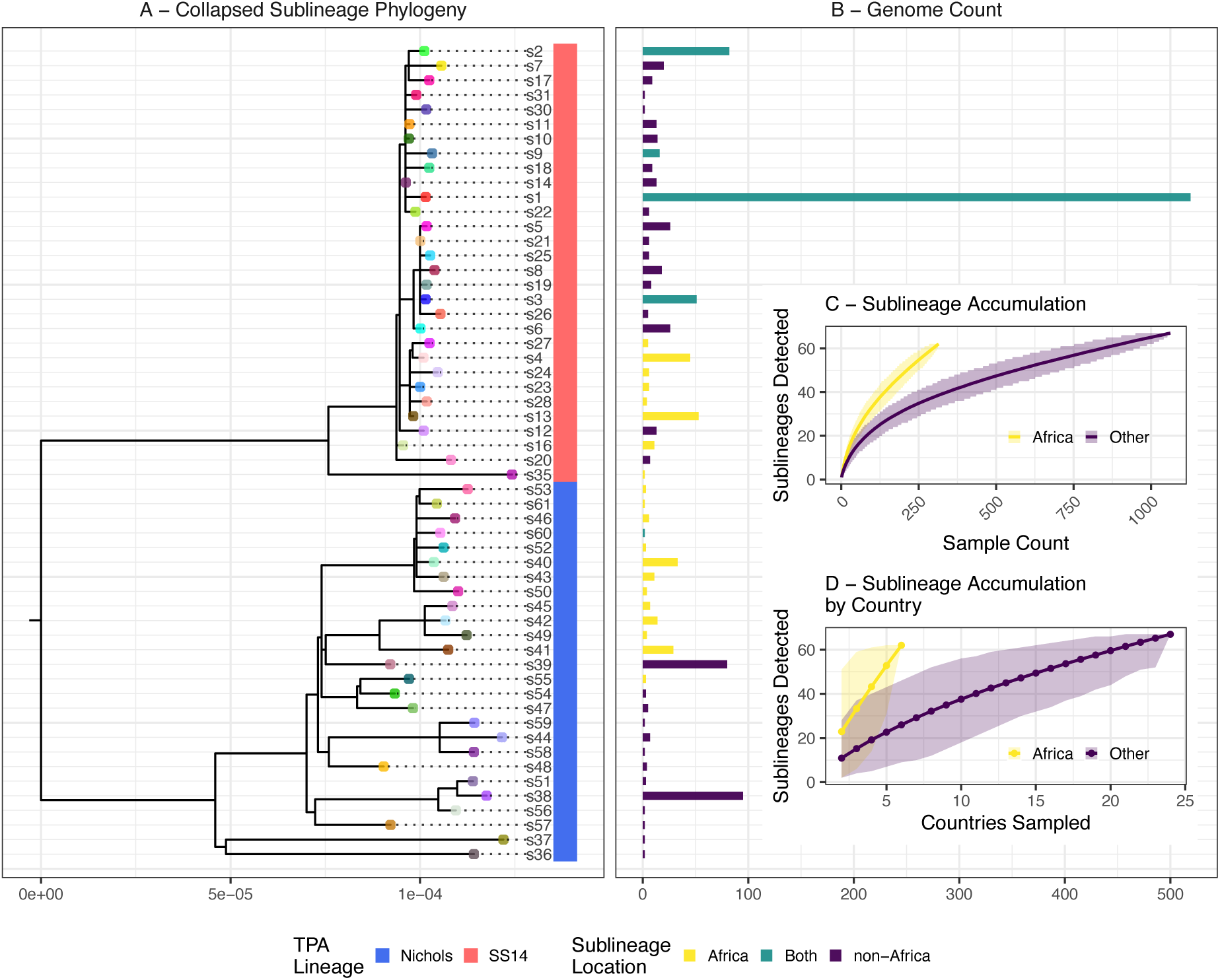
African and global sublineages are distinct. A - Collapsed whole genome phylogeny showing one tip per sublineage (African genomes only). B – Genome count per sublineage. Sublineages are classified based on being exclusive to Africa, absent from Africa, or present globally and in Africa. C - Sublineage accumulation inside and out of Africa with increased sampling. D – Sublineage accumulation as more countries are sampled. Whilst the curve for the 21 non-African countries is starting to plateau, new African sublineages are still being discovered at a steep rate after only 6 countries sampled. Lines show median estimated sublineage count, shading shows 2.5%-97.5% quantiles from 10,000 permutations. Analysis for C and D includes singleton sublineages.

### Sublineage dissemination within and between African countries

The 314 African genomes were distributed across 25 sublineages, but the majority of these sublineages were grouped into one of four larger phylogenetic clades (Figure 3, Supplementary Figure 5). To understand local sublineage dynamics we focussed on South Africa, where we had more representative sampling - all provinces over a 16-year timespan (2007-2023). As a counterpoint we compared these data with our previously published data from the UK^7^. We found a diverse collection of sublineages circulating within South African provinces, but the proportion of sublineages varied by province (Supplementary Figure 11). For example, sublineage 2 was most prevalent in KwaZulu-Natal and less common elsewhere. Moreover, no single province contained all the sublineages seen in South Africa, a pattern that replicated the UK^7^ (Supplementary Figure 12). Metropolitan provinces (e.g. Gauteng, which includes Johannesburg; London in the UK) had the highest number of sublineages (Supplementary Figure 12). However, when we used rarefaction to adjust for sampling, sublineage richness was not elevated in Gauteng compared to other South African provinces. From our accumulation analysis, we estimate that at least three geographically distinct sampling locations are needed to capture ∼50% of novel diversity in a new country (mean range 18.7-93.8% from 10,000 permutations, Supplementary Figure 12D).

**Figure 3.**
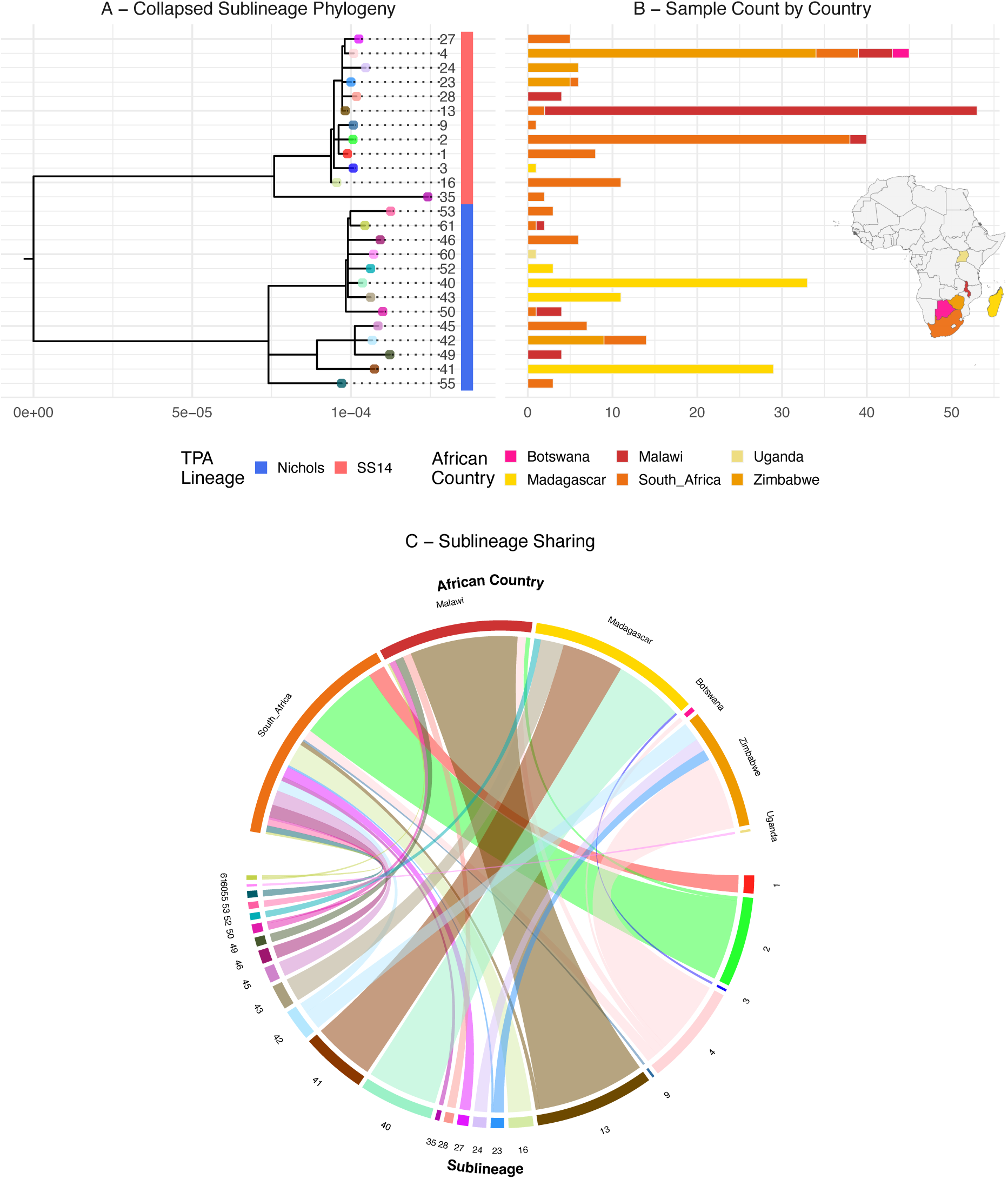
Sharing of sublineages between African countries is uncommon. A - Whole genome of African TPA genomes, showing a single representative tip for each sublineage. Coloured track indicates Lineage. B – Sample count and country of sampling. C – Chord diagram of sublineage sharing between countries.

Of the African sublineages, 18/25 were restricted to a single country and 6/25 were found in two African countries (Supplementary Figure 14). The latter included sublineage 2 described above (South Africa, n=38; Malawi, n=2), which was the only sublineage found globally and in multiple African countries.

Amongst the twenty sublineages specific to Africa, sublineage 13 was detected in South Africa in 2016 (2/53) and in Malawi in 2020-2021 (51/53). Similarly, a sublineage 50 genome was identified in South Africa in 2016 (1/4) and in Malawi in 2020-2021 (3/4), whilst sublineage 61 was detected in South Africa in 2009 (1/2) and then in Malawi in 2021 (1/2). We also found sharing between South Africa and Zimbabwe, with samples from sublineage 23 detected in Zimbabwe in 2015 and 2023 (5/6) and in South Africa in 2021 (1/6). Sublineage 42 was found to be persistently circulating in both South Africa (2011-2021, 5/14) and Zimbabwe (2015-2023, 9/14). Finally, sublineage 4 was found in four African countries, having been first detected in South Africa (2009-2023; 5/45), and was also detected in Zimbabwe (2015, 2022-2023; 34/45), Malawi (2020; 4/45) and Botswana (2023; 2/45). Notably, samples from the different countries were polyphyletic in the sublineage 4 phylogeny (Supplementary Figure 14), suggesting ongoing circulation of this sublineage amongst Southern African countries. We examined the metadata for patients infected with sublineage 4 and found a mix of women (48.9%, 22/45), men (33.3%, 15/45) and unrecorded (17.8%, 8/45), implying predominantly heterosexual transmission.

We linked patient gender to 70.8% (975/1376) of genomes across the global dataset (Supplementary Figure 15). Of these, 60.8% (837/1376) were from men and 10.0% (138/1376) from women, consistent with enriched sampling of GBMSM in HIC syphilis epidemics. Gender data were available for 67.8% (213/314) of African genomes (Madagascar data were unavailable). In Zimbabwe, 50.9% (28/55) of genomes came from women, suggesting we had sampled from heterosexual transmission networks. Indeed, only one of 104 African patients disclosing their sexual orientation identified as GBMSM (a bisexual infected with a sublineage 2 strain). Consistent with global patterns, in South Africa the majority of globally circulating sublineage 1 (75%, 6/8), 2 (89.8%, 33/38) and 9 (1/1) samples came from men, and sublineages 1 and 9 were previously associated with GBMSM networks in a European study^7^. However, only 1/102 South African individuals identified as GBMSM.

### Role of sublineage dissemination in antimicrobial resistance

Consistent with widespread increases in macrolide-resistant syphilis, 68.6% (944/1376) of genomes carried either one of two macrolide resistance-conferring alleles, A2058G (66.6%, 917/1376) or 2059G (2.0%, 27/1376). However, excluding South Africa, where resistance rates were high (41.2%, 42/102), only 9.9% (21/212) of TPA from the other African countries were predicted to be macrolide resistant (Figure 4A). Specifically, 50% (1/2) of Botswana genomes, 2.5% (2/80) of Malawi genomes, 12.2% (9/74) of Madagascar genomes and 16.4% (9/55) of Zimbabwe genomes were predicted to be resistant, whilst the only Uganda genome was predicted to be sensitive.

**Figure 4.**
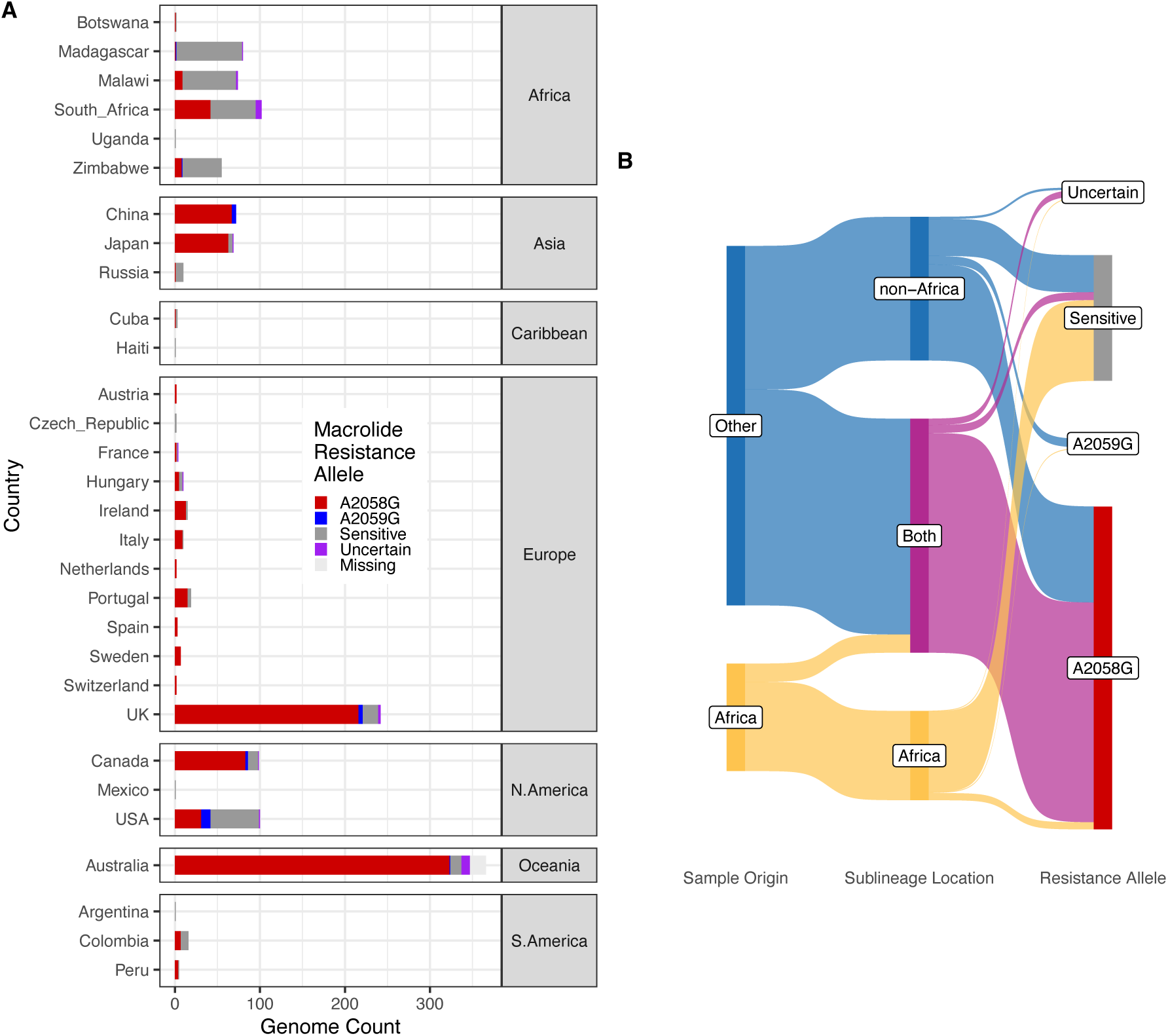
Macrolide resistant rates in syphilis from Africa are substantially lower than globally. A - Presence of macrolide resistance alleles (A2058G, A2059G) and wildtype by Country. Uncertain = mixed alleles at position. Note that samples from USA are older, and in some cases predate the emergence and spread of macrolide resistance in the early 2000s^5^. B – Sankey diagram of Sample Origin (Africa or elsewhere), Sublineage type (African, non-African, Both) and Macrolide Resistance Allele.

We examined the distribution of macrolide resistance alleles at the sublineage level, considering whether sublineages were exclusive to Africa, absent from Africa, or present in both Africa and elsewhere (Supplementary Figure 16). Whilst 20/31 non-African sublineages contained genomes predicted to be macrolide resistant, only 5/20 African-only sublineages did. Moreover, 4/5 globally shared sublineages contained genomes predicted to be resistant, dominated by the major global sublineage 1 (519/1376 genomes in the total dataset), of which 94.8% (492/519) samples were predicted to be resistant. Notably, whilst these globally shared or imported sublineages accounted for only 16.2% (51/314) of African genomes, they accounted for 63.5% (40/63) of the macrolide resistance (Figure 4B).

In South Africa, no genotypic resistance (0/20 genomes) was observed in genomes collected between 2007-2012. The A2058G allele was first seen in a sublineage 2 genome in 2013^15^, with A2058G subsequently detected in 51.2% (42/82) of all South African genomes for the period of 2013-2023. This is consistent with the influx of globally shared macrolide resistant sublineages 1 and 2 into South Africa from 2013 onwards, representing 7.8% (8/102) and 37.3% (38/102) of South African genomes respectively, and together accounting for 88.1% (37/42) of resistant syphilis in South Africa (Supplementary Figure 17). The remaining resistant syphilis was attributed to sublineage 16, found exclusively in South Africa in our dataset, of which 45.5% (5/11) of genomes were predicted to be resistant. The sublineage 16 subtree (Supplementary Figure 18) indicates resistance occurs in only one of two clades, with the earliest resistant genome sample collected in 2018 in Free State and 4/5 resistant genomes subsequently found in KwaZulu Natal.

## Discussion

Due to the paucity of syphilis genome data from Africa we established prospective surveillance sites in four African countries. In doing so, we provide the first TPA genomes from Botswana and Uganda, whilst substantially increasing the number of genomes from South Africa and Zimbabwe. By integrating TPA genome data from other sites including Madagascar and Malawi^6,10^, we provide the most comprehensive genomic analysis of African syphilis to date.

We show the proportion of Nichols- and SS14-lineages circulating in most African countries is approximately 20:80. This is consistent with previous global sampling^5,6^, despite African diversity being made up of a different and more diverse group of sublineages. In contrast to the picture amongst HICs, where global sharing of sublineages is extremely common^5^, the majority of African genomes represented novel diversity and sublineages that were unique to Africa. Despite unevenly sampling from just six African countries, our accumulation analysis indicates that we have not yet approached a plateau in terms of identifying novel diversity in Africa.

We found a high number of novel sublineages despite limited genomic sharing between African countries, and we speculate this is explained by founding effects and local diversification. This pattern extended to our analysis of regional diversity in South Africa, where different provinces were dominated by different sublineages, possibly reflective of lower population mobility compared to Europe and North America. However, several African sublineages share common ancestral nodes dating back to the 1980s, and at least one sublineage specific to Africa (sublineage 4) was circulating widely in four countries and amongst both men and women – likely reflecting a large regional transmission network.

Within countries, no single South African Province captured all circulating diversity, and we estimate that at least three distinct sampling locations are needed to capture ∼50% of novel diversity in a new country. Where resources are scarce, targeting metropolitan areas where diversity is high and elevated case numbers make sampling more straightforward. Despite recent efforts here and elsewhere^10^ to prospectively collect and sequence African syphilis, most African genomes in our study derive from Southern Africa, and additional sampling focussed on West, Central and East Africa is necessary.

Against this background of discrete local transmission of predominantly macrolide-sensitive African sublineages, we observed multiple independent introductions of macrolide-resistant, globally circulating sublineages^5,9^. These introductions were most common in South Africa, where sublineage 1 was independently introduced at least four times, alongside introductions of global sublineages 2 and 9. In contrast, we found only two phylogenetically related sublineage 2 genomes in Malawi, and a single sublineage 3 genome in Madagascar, with no global sublineages detected in Zimbabwe or Botswana. Strikingly, in the 10 years following first detection, these imported macrolide-resistant global sublineages increased in prevalence to account for half of TPA genomes in South Africa, suggesting either a fitness advantage or a strong network effect. Our data demonstrate how rapidly a sexually transmitted pathogen can spread within a population and have implications for the use of global genomic AMR data to inform local treatment guidelines in the African region. Similar trends have been seen with global transmission of multi-drug resistant *Neisseria gonorrhoeae*^18,19^. In contrast to rising macrolide resistance in South Africa, and similar to a previous study^15^, our contemporaneous sampling in neighbouring Zimbabwe (2015, 2022-2023) found only 16.4% resistance and no imported global lineages. This may be because the majority of global sublineages sampled in South Africa were found only in men, which may reflect differences in care seeking behaviour, culture and believe systems, or inadvertent sampling or bridging from GBMSM networks. Transmission dynamics of STIs are highly sensitive to network structure^20^ which can influence transmission of a pathogen or specific lineage. Our observation that this has been primarily observed in South Africa may reflect the country’s greater international connectivity, differences in sexual network dynamics, and more extensive surveillance and sampling compared with neighbouring countries.

We identified a novel sublineage 16 which was distinct to South Africa, and in which macrolide resistance may be emerging independent of global importation. *De novo* selection of macrolide resistance may reflect both on- and off-target treatment interventions. Macrolides are no longer recommended treatment for syphilis in most HICs, but in some cases are used in LMICs, and are still recommended by WHO as an alternative therapy to BPG^21,22^. Moreover, macrolides are used for treating other infections suggesting bystander selection is likely^23^. Whilst macrolide resistant syphilis remains less common in Africa than globally, our data show it is now emerging rapidly in some countries, suggesting that azithromycin should no longer form part of WHO recommendations for syphilis treatment.

Use of syndromic management for STIs in most LMICs has significantly impeded monitoring of transmission and AMR. Our data demonstrate the value of high-resolution genomic data for surveillance and to inform interventions, including global recommendations on antimicrobial usage and vaccines. Development of appropriate regional surveillance systems in Africa could potentially be modelled off similar strategies for gonorrhoea^24^. The need for such systems will become more pressing as AMR continues to rise and novel strategies such as doxycycline post-exposure prophylaxis are rolled out in both high and low-income settings. African TPA sublineages appear relatively stable and geographically restricted. Future surveillance efforts should consider the opportunities to target different groups, for example antenatal and key populations, to ensure we accurately reflect transmission chains and genomic diversity in populations at most risk. Treating cases or populations confirmed to carry local lineages with different antimicrobial regimens, alongside global efforts to develop candidate TPA vaccines using data linked to the smaller number of dominant macrolide resistant global lineages, could be efficacious. Either way, with ever increasing globalisation and population migrations, ignoring the greater diversity in African TPA would be a mistake.

## Data Availability

Novel sequencing data are available from the European Nucleotide Archive under BioProject accessions PRJEB60271 and PRJEB63120. Additional sequences are listed in Supplementary Data 1, listing ENA/SRA or NCBI accessions. All metadata and key intermediate files along with fully worked analysis code are available at https://github.com/matbeale/MAGUS_analysis, along with any additional custom scripts used.

https://www.ebi.ac.uk/ena/browser/view/PRJEB60271

https://www.ebi.ac.uk/ena/browser/view/PRJEB63120

## Data & Code Availability

Novel sequencing data are available from the European Nucleotide Archive under BioProject accessions PRJEB60271 and PRJEB63120. Additional sequences are listed in Supplementary Data 1, listing ENA/SRA or NCBI accessions. All metadata and key intermediate files along with fully worked analysis code are available at https://github.com/matbeale/MAGUS_analysis, along with any additional custom scripts used. No patient identifiable information is contained in the manuscript or supplementary information.

## Author Contributions

Conceptualisation: MAB, MM, NRT; Methodology: MAB, SB, RH, MM; Investigation: MAB, SB, RH, KEA; Resources: MAB, YAS, CM, ENJ, EM, RAF, MM, NRT; Data Curation: MAB, MM, RH, SB, KEA, ED, AM, BB, SM, MO, DPK, RK, EK, DK, MPM, JMEV, BDCD; Formal Analysis: MAB; Visualisation: MAB; Supervision: NRT; Project Administration: MAB, MM, CM, YAS, ENJ, EM, RF, NRT; Writing – Original Draft: MAB; Writing – Review and Revision: All Authors.

## Acknowledgements

The authors thank all participants of the MAGUS study and members of the research teams, as well as additional technical staff at NICD for the South Africa samples. The authors acknowledge support at the Wellcome Sanger Institute from the Core Sequencing, Parasites and Microbes (PAM) Informatics, PAM Samples, and PAM Core Laboratory team for assistance with sample governance, processing, sequencing and data management. We thank M. Taouk and D. Williamson for sharing unpublished pseudonymised metadata for Australian patients^14^, and F. Aghakhanian and J. Parr for sharing unpublished metadata from Malawian, Chinese and Colombian patients^10^.

This work was part-funded by a Gates Foundation grant (INV-035896) to NRT, MAB, MM, CM, YAS, ENJ, and RF. MAB, KA and NRT were supported by Wellcome funding to the Sanger Institute (206545/Z/17/Z; ‘Wellcome Sanger Institute Quinquennial Review 2021-2026’). MM is funded by the National Institute for Health and Care Research (NIHR304258).

This work was supported, in whole or in part, by the Gates Foundation [INV-035896] and [206545/Z/17/Z] ‘Wellcome Sanger Institute Quinquennial Review 2021-2026’. The conclusions and opinions expressed in this work are those of the authors alone and shall not be attributed to the Gates Foundation, the Wellcome Trust, NIHR or the Department of Health and Social Care. The funders had no role in study design, data collection and analysis, decision to publish or preparation of the manuscript. Under the grant conditions of the Foundation, a Creative Commons Attribution 4.0 License has already been assigned to the Author Accepted Manuscript version that might arise from this submission. Please note works submitted as a preprint have not undergone a peer review process.

