## Supplementary Methods for "High diversity amongst African *Treponema pallidum* genomes provides a window into global transmission dynamics of syphilis: A genomic epidemiology study"

### Sequencing

Metagenomic swab DNA samples were subjected to library prep using the pooled sequence capture method described in detail previously[1,2]. Extracted genomic DNA was sheared to 100-400 bp using an LE220 ultrasonicator (Covaris Inc.). Libraries were prepared (NEBNext Ultra II DNA Library prep Kit, New England Biolabs, Massachusetts, USA) using initial adaptor ligation and barcoding with unique dual-indexed barcodes (Integrated DNA Technologies, Iowa, USA). Dual-indexed samples were amplified (6 cycles of PCR, KAPA HiFi kit, Roche, Basel, Switzerland), quantified (Accuclear dsDNA Quantitation Solution, Biotium, California, USA), then pooled in preassigned groups of between 20 and 32 (depending on batch size at time of sequencing) to generate equimolar pools based on Total DNA concentration. Pools were assigned based on similar qPCR Cq, with outlying low Cq samples diluted to be similar to other samples in the pool to prevent them dominating sequencing reads. 500 ng pooled DNA was hybridised using 120-mer RNA baits (SureSelect XT Target enrichment system, Agilent technologies; Bait design ELID ID 0616571). Enriched libraries were sequenced on NovaSeq 6000 on SP flowcells (Illumina Inc., San Diego), to generate 150 bp paired end reads.

### Bioinformatics and Phylogenetics

We used Kraken[3] v2.1.2 to filter and extract Treponema-specific reads, followed by trimming with Trimmomatic[4] v0.39 and downsampling to a maximum of 3,500,000 using seqtk v1.0 (available at https://github.com/lh3/seqtk) as previously described[1]. Novel genomes from Africa were contextualised using a dataset comprising published genomes from previous studies[1,5–17], comprising all high quality TPA genomes publicly available in August 2024, with reads processed in the same way (except using a cut-off of 2,500,000 reads, as previously applied).

For phylogenetic analysis, we used an established pipeline to map *Treponema*-specific reads to a custom version of the SS14_v2 reference genome (NC_021508.1), after first masking 12 repetitive Tpr genes (Tpr A-L), two highly repetitive genes (arp, TPANIC_0470), and five FadL homologs (TPANIC_0548, TPANIC_0856, TPANIC_0858, TPANIC_0859, TPANIC_0865) using bedtools v2.29 maskfasta as described previously[5]. Reads were mapped using BWA mem v0.7.17, followed by indel realignment (GATK Indel Realigner v3.7), optical duplicate marking (Picard MarkDuplicates v1.126), and variant calling (samtools v1.6 and bcftools[18] v1.6). We required a minimum of five supporting reads (two per strand) to call a variant, and a variant frequency/mapping quality cut-off of 0.8. Sites not meeting these criteria were masked to 'N' in the final pseudosequence. We repeated the masking of these 19 genes on the final multiple sequence alignment using remove_block_from_aln.py available at https://github.com/sanger-pathogens/remove_blocks_from_aln/ to ensure sites originally masked in the reference were not inadvertently called with SNPs. Masking these regions accounted for 30,071 reference sites (2.7% of the total genome length).

After pseudosequence generation, we excluded genomes for which ≥25% of reference positions failed our strict filtering criteria. Combined with 1229 contextual genomes, this left us with a total dataset comprising 1376 whole genomes. We used Gubbins[19] v3.2.1 to identify and mask additional recombination in our multiple sequence alignment, before generating a SNP-only alignment containing 1,892 variable sites using snp-sites[20] v2.5.1, and using IQ-Tree[21] v1.5.12 to infer a GTR+F mode, inputting invariant sites using the `-fconst` argument and requiring 1,000 UltraFast bootstraps[22].

For phylogenetic clustering, we performed joint ancestral reconstruction of SNPs in the maximum likelihood phylogeny using pyjar (available at https://github.com/simonrharris/pyjar), and then used rPinecone[23] (available at https://github.com/alexwailan/rpinecone) with a 10 SNP threshold as previously.

For temporal analysis, we initially evaluated the temporal signal in our full 1376 genome maximum likelihood phylogeny. This revealed that although the SS14-lineage shows a robust temporal signal, this was not the case for the Nichols-lineage. We therefore proceeded with temporal analysis only for SS14-lineage. To representatively subsample the 1376 genome dataset whilst maintaining genetic diversity, we used treemer[24] v0.3, which iteratively prunes tips from a phylogeny based on nearest neighbour relationships. We pruned the 1376 genome phylogeny to 600 genomes, removing 25 genomes per iteration. Of these 440/600 genomes were SS14-lineage, and the temporal signal was maintained; we used these genomes for temporal analysis. We initially ran BEAST1[25] v1.10.4 in triplicate on our recombination-masked SNP-only alignment containing 440 genomes, correcting for invariant sites using the 'constantPatterns' argument. We evaluated both Strick Clock model[26] (starting rate prior 1.78 $x10^{-7}$) and an Uncorrelated Relaxed Clock model[27], with a GTR substitution model[28] and diffuse gamma distribution prior (shame 0.001, scale 1000) over 500 million Markov Chain Monte Carlo (MCMC) cycles with a 50 million cycle burnin, using BEAGLE libraries to enable analysis with fast Graphical Processing Units[37]. We evaluated constant, relaxed lognormal, exponential and Bayesian Skyline population distributions[29]. All MCMC chains converged with high effective sample sizes (ESS). We evaluated the marginal distribution of `ucld.stdev' and this favoured a relaxed clock model. We then evaluated the optimal model using Path Sampling and Stepping Stone analysis[30,31], determining that the Relaxed Lognormal model with Bayesian Skyline was optimal. This tree inferred a molecular clock of 1.75 $x10^{-7}$ substitutions per site per year, consistent with other recent analyses of contemporary *T. pallidum* evolution[1,5,7,8], but faster than those incorporating ancient genomes[32–34]. We repeated the optimal model, implementing a continuous time Markov model by adding Continent information as a trait partition, and enabling ancestral state reconstruction and state change rate tracking[26,35]. We also added parameters for counting state changes (Markov jumps) and time between state changes (Markov rewards)[35]. These MCMC chains were run for 1 billion cycles, and all parameters converged with high ESS. We used logcombiner v2.6.3 with a 20% burnin to generate consensus log and tree files from the three independent runs (resampling every 3 million states), and treeannotator v1.8.4 to create a median maximum credibility tree.

Since we were unable to infer a time-scaled tree for the full 1376 genome dataset, to make inferences about ancestral phylogeography across all samples we used the map.simmap command in phytools[36] v2.4.4 with 100 simulations to implement a continuous time Markov model on the recombination filtered maximum likelihood phylogeny from IQ-Tree, using Continent as a trait. Phylogenetic clades close to the tips in the maximum likelihood tree are very clonal, and in some cases we had zero-distance branch lengths and polytomies; to resolve this we added a nominal value (1e-6) to branches with zero distance and forced bifurcation using the multi2di command in ape[37] v5.8.1.

We generated subtrees by inferring the most recent common ancestor using the getMRCA function in ape and then using the `tree_subset` function in ggtree. We created representative subtrees with a single tip per sublineage using the makeCollapsedTree function in treespace[38] v1.1.4.3.

For evaluating accumulation of sublineages, we sampled genomes sequentially in R and counted sublineage accumulation (including singletons), evaluated over 10,000 permutations. To calculate sublineage accumulation by country or region, we used the `poolaccum` command in vegan[39] v2.6.6.1, using sublineage counts (including singletons) in place of taxon counts, calculated over 10,000 permutations. For evaluating richness of sublineages in different regions/countries, we performed rarefaction using iNEXT[40] v3.0.2.

Macrolide resistance SNPs were inferred from enriched metagenomic data using the stringent competitive mapping approach described previously[1].

For spatial mapping, we downloaded publicly available shape files with province boundaries for South Africa from Humanitarian Data Exchange (https://data.humdata.org/dataset/cod-ab-zaf). We also downloaded shape files for Africa from the ICPAC Geoportal (available at https://geoportal.icpac.net/layers/geonode%3Aafr_g2014_2013_0). We used the sf[41] v1.0.16, broom[42] v0.7.10 and ggplot2 packages to process and plot map data, and the scatterpie[43] v0.1.7 package to plot pie charts. Chord diagrams were generated using the circlize[44] v0.4.16 package and sankey diagrams were generated using ggsankey[45] v0.0.99999. All phylogenetic trees were plotted in R using ggtree[46] v2.5.1. All figures used ggplot2[47] for plotting and cowplot[48] v1.1.3 for arranging panels.

### Analysis of Patient Gender

Basic demographic metadata was collected for patients recruited into the Multi-country Aetiology of Genital Ulcer Survey (MAGUS), and this was also available for samples collected from South Africa by NICD. We collated further patient gender metadata from published studies linked to genomes. Two large studies[6,8] of *T. pallidum* genomes did not contain information on patient gender, and we approached the authors of these studies for information on samples from Colombia, China, Malawi and Australia.

Data from Australia was collected through Public Health Laboratories from populations in which heterosexuals were uncommon; to prevent deductive disclosure the data cannot be fully linked to genome identifiers in published material. Rather than include this information in Supplementary Data 1, we therefore used this data to calculate gender proportions according to country and sublineage, and this information was then integrated with calculations for other global genomes.

Per sample gender data was unavailable for Madagascar genomes, but in the original study 61% of patients recruited into the original study were male, with 98% of individuals identifying as heterosexual, whilst those in which *T. pallidum* DNA was detected were predominantly male (79%)[49] – a subset of these were sequenced in a previous study[7].

### Supplementary Figures

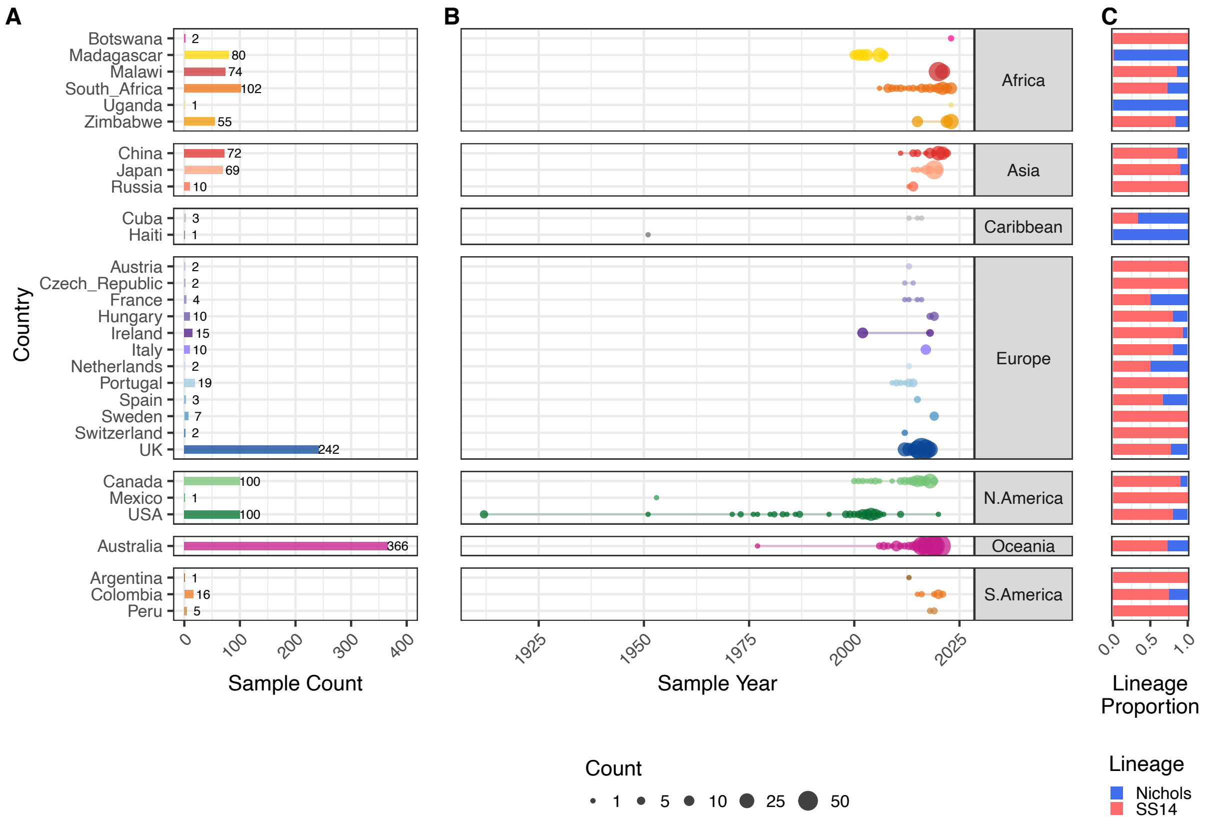


**Supplementary Figure 1.** Distribution of 1376 *T. pallidum* subspecies *pallidum* genomes by country and sample collection year.

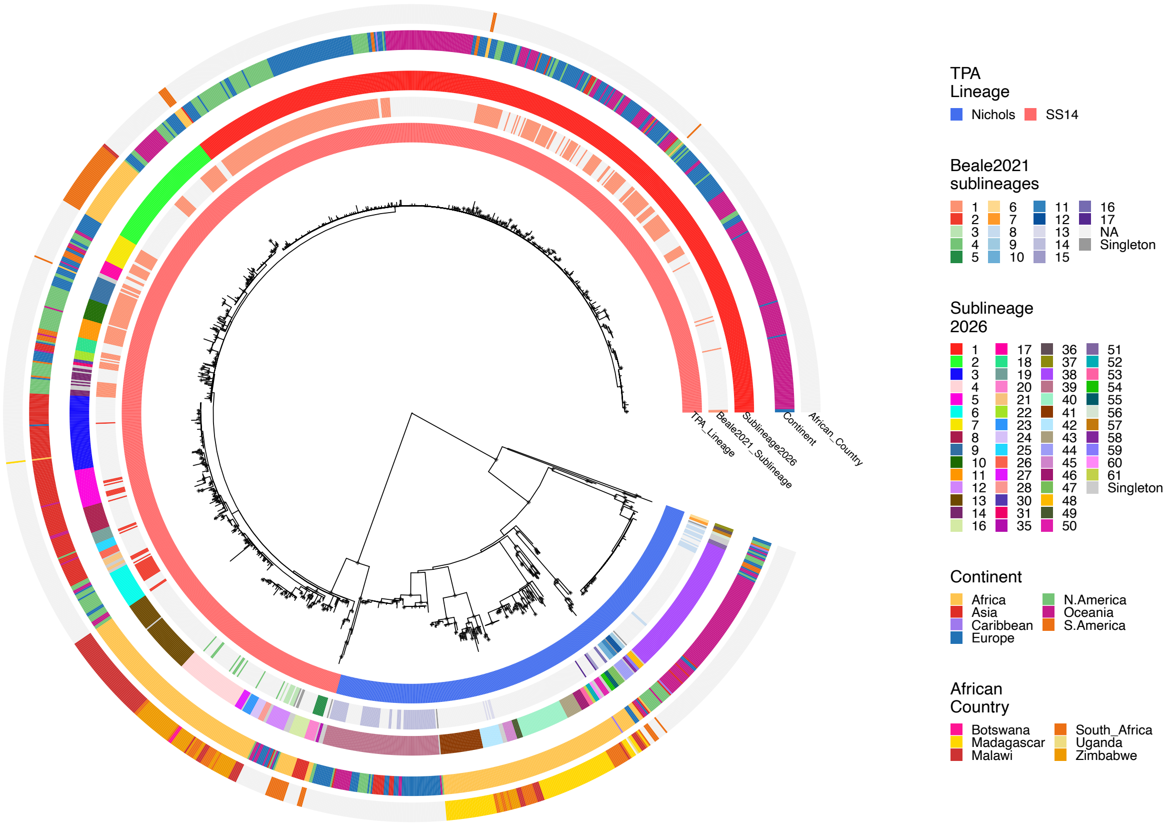


**Supplementary Figure 2.** Maximum likelihood whole genome phylogeny of 1376 *T. pallidum* subspecies *pallidum* genomes. Coloured tracks indicate Lineage, Sublineage previously defined in Beale 2021 (pale grey if genomes were not available/classified in that study), Sublineages redefined for the current dataset, Continent, African country (where sample is from Africa).

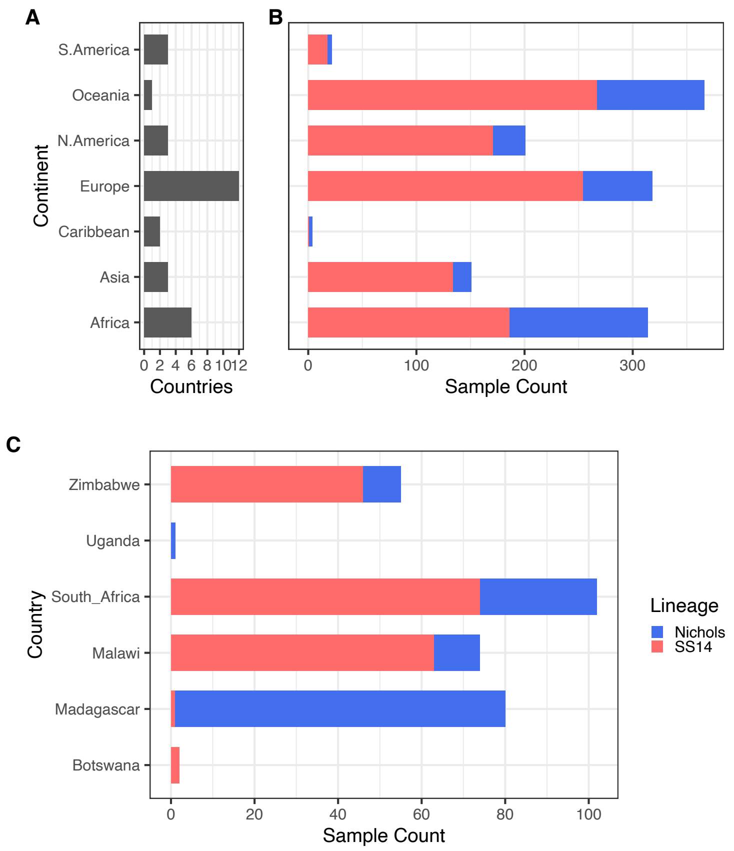


**Supplementary Figure 3.** Global distribution of major TPA lineages. A – Number of countries per continent with TPA genomes. B – Total genomes per continent coloured by SS14- and Nichols- lineages. C – Total genomes within African countries, coloured by SS14 and Nichols lineages.

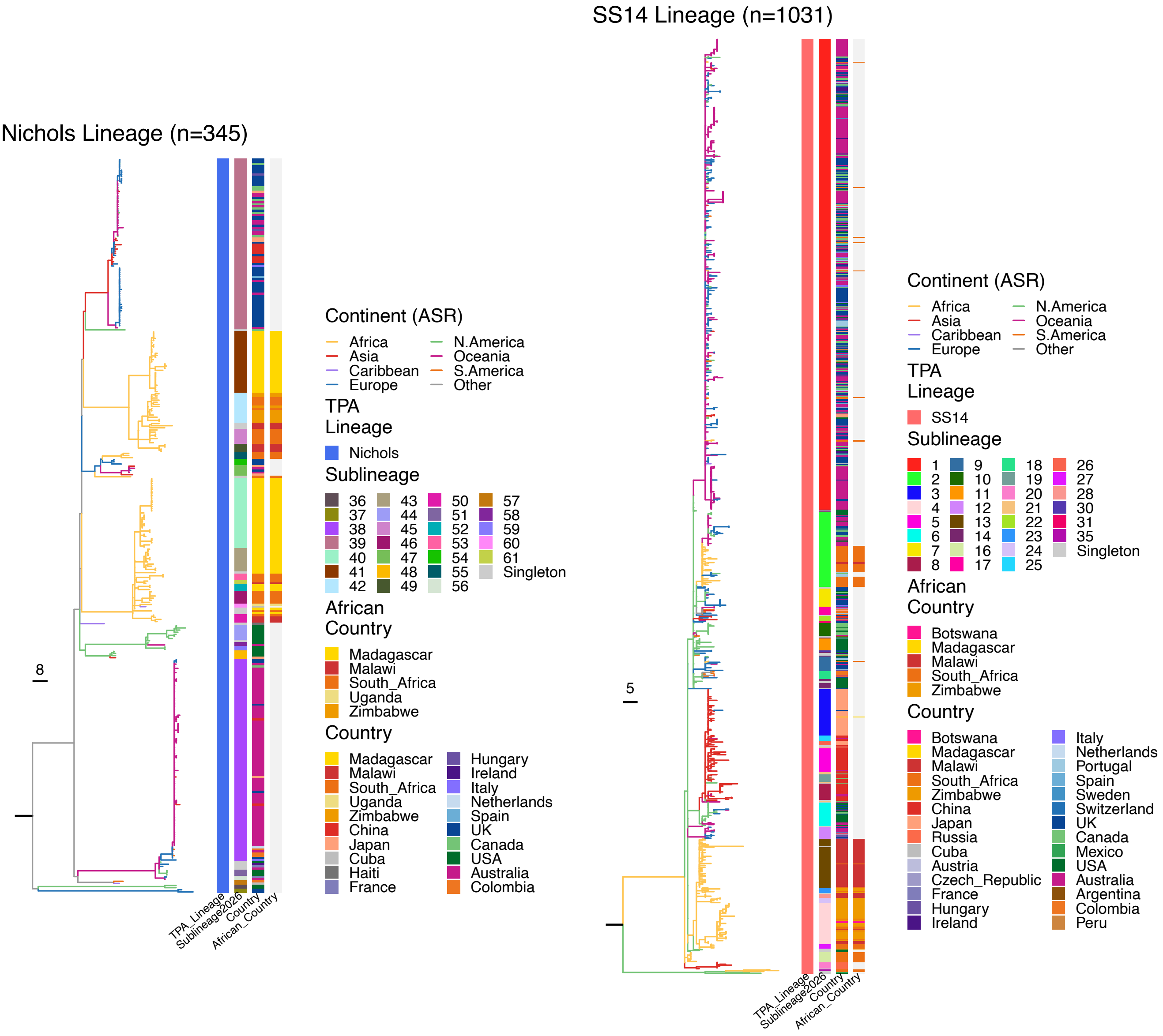


**Supplementary Figure 4.** Maximum likelihood whole genome phylogenies of Nichols- and SS14-lineages, separated for clarity. Branches are coloured based on ancestral state reconstruction of Continental location, and branch lengths are scaled by substitutions. Coloured tracks indicate Lineage, Sublineage, Continent, Country, and African Country (where sample is from Africa).

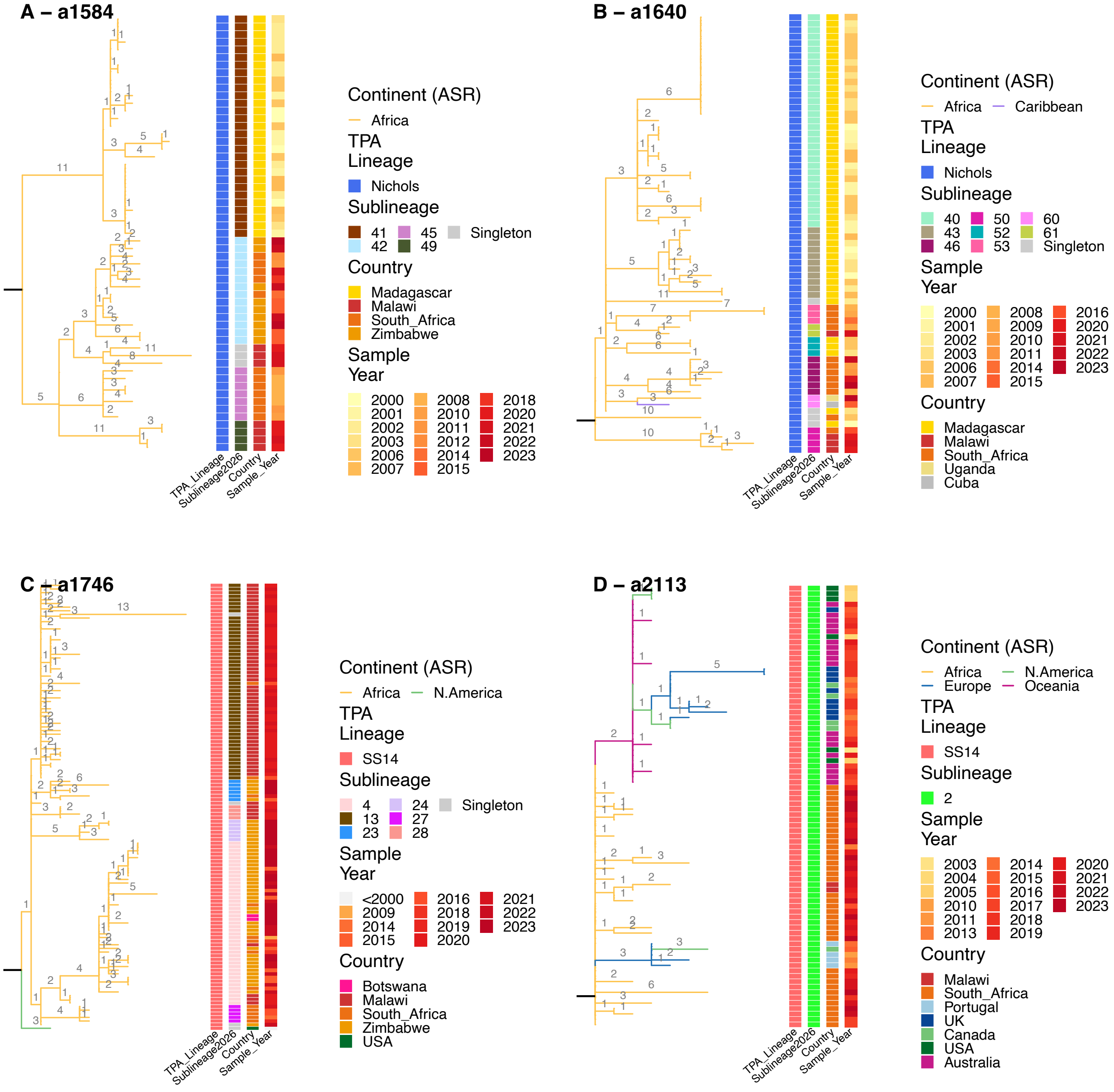


**Supplementary Figure 5.** Maximum likelihood subtrees descending from major common ancestral nodes (a2584, a1640, a1746, a2113) occurring in Africa (comprising >2 African genomes and excluding the global and polyphyletic sublineage 1). Branches are coloured based on ancestral reconstruction of continental state and scaled according to substitutions (indicated by numbers). Coloured tracks indicate Lineage, Sublineage, Country, and Year of sampling.

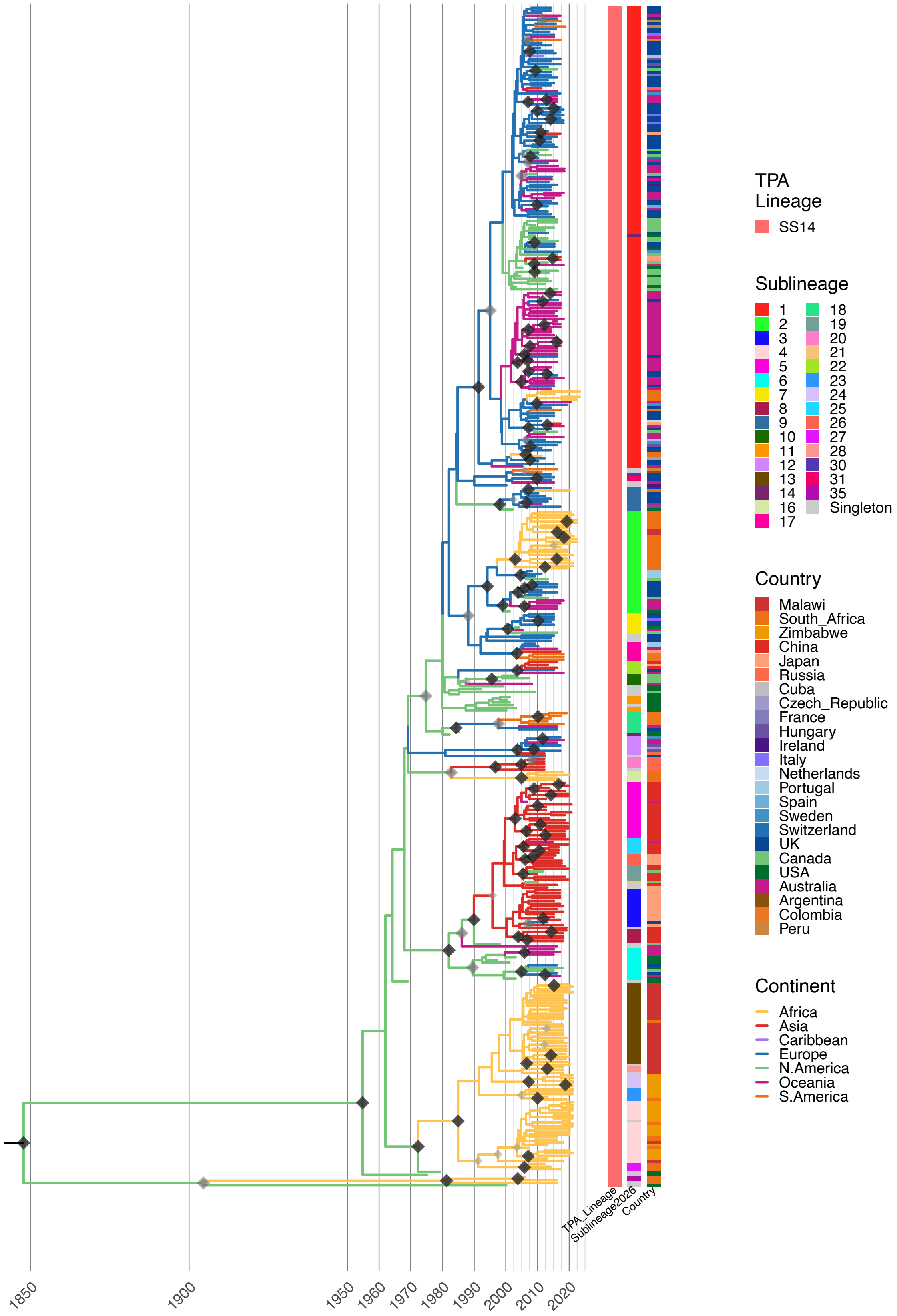


**Supplementary Figure 6.** Time-scaled maximum credibility phylogeny of 440 SS14-lineage whole TPA genomes. Branches are coloured according to the inferred location (Continent) at each ancestral node. Diamond points indicate posterior likelihood (>0.8 light grey, >0.91 dark grey, >0.96 black). Coloured tracks indicate TPA Lineage, Sublineage and Country.

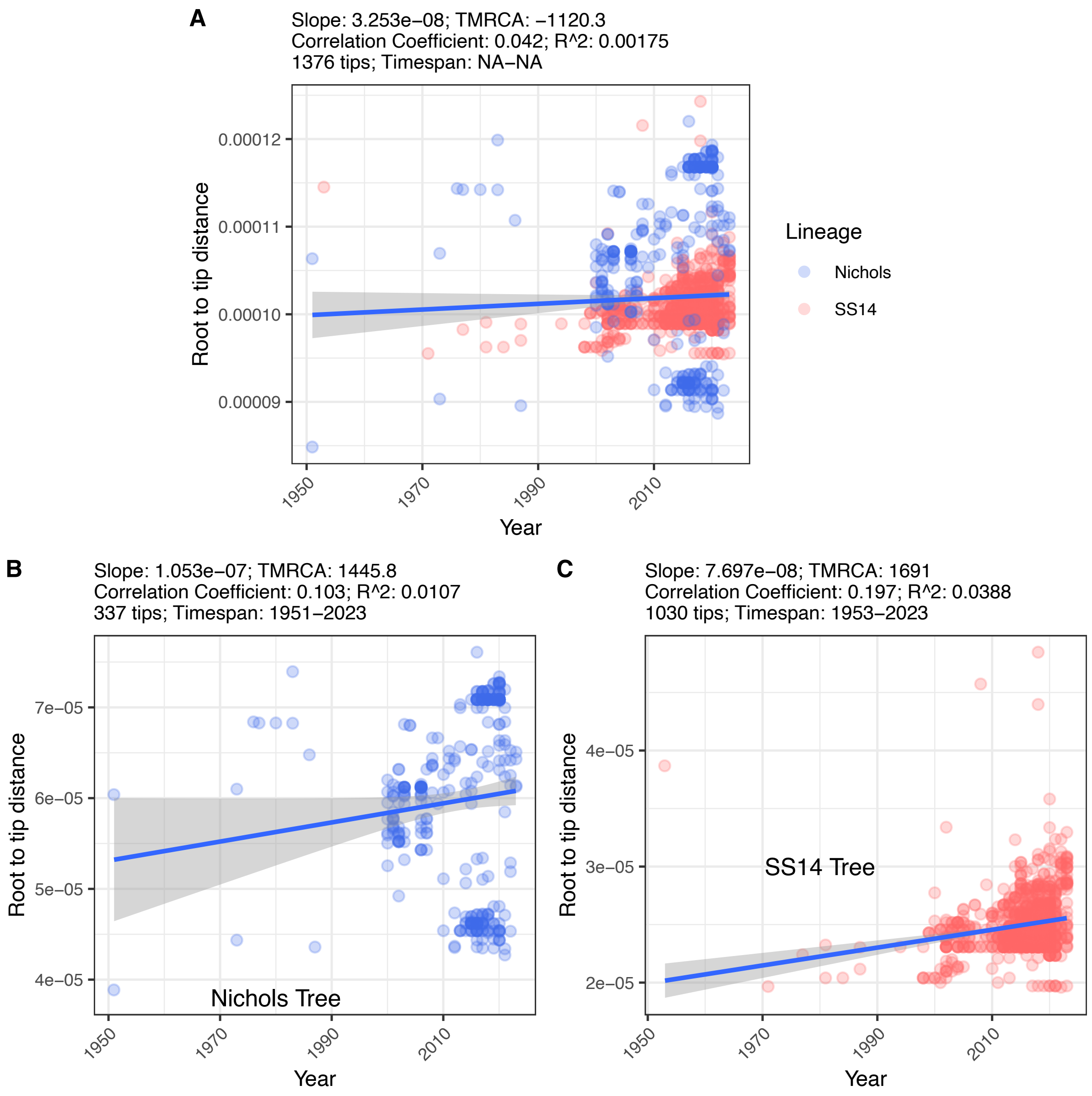


**Supplementary Figure 7. Nichols-lineage lacks a robust linear temporal signal in this dataset and was unsuitable for phylodynamic analysis using BEAST.** Root-to-tip corelation analysis of full 1376 genome dataset (A), and partitioned to show Nichols-lineage (B) and SS14-Lineage (C).

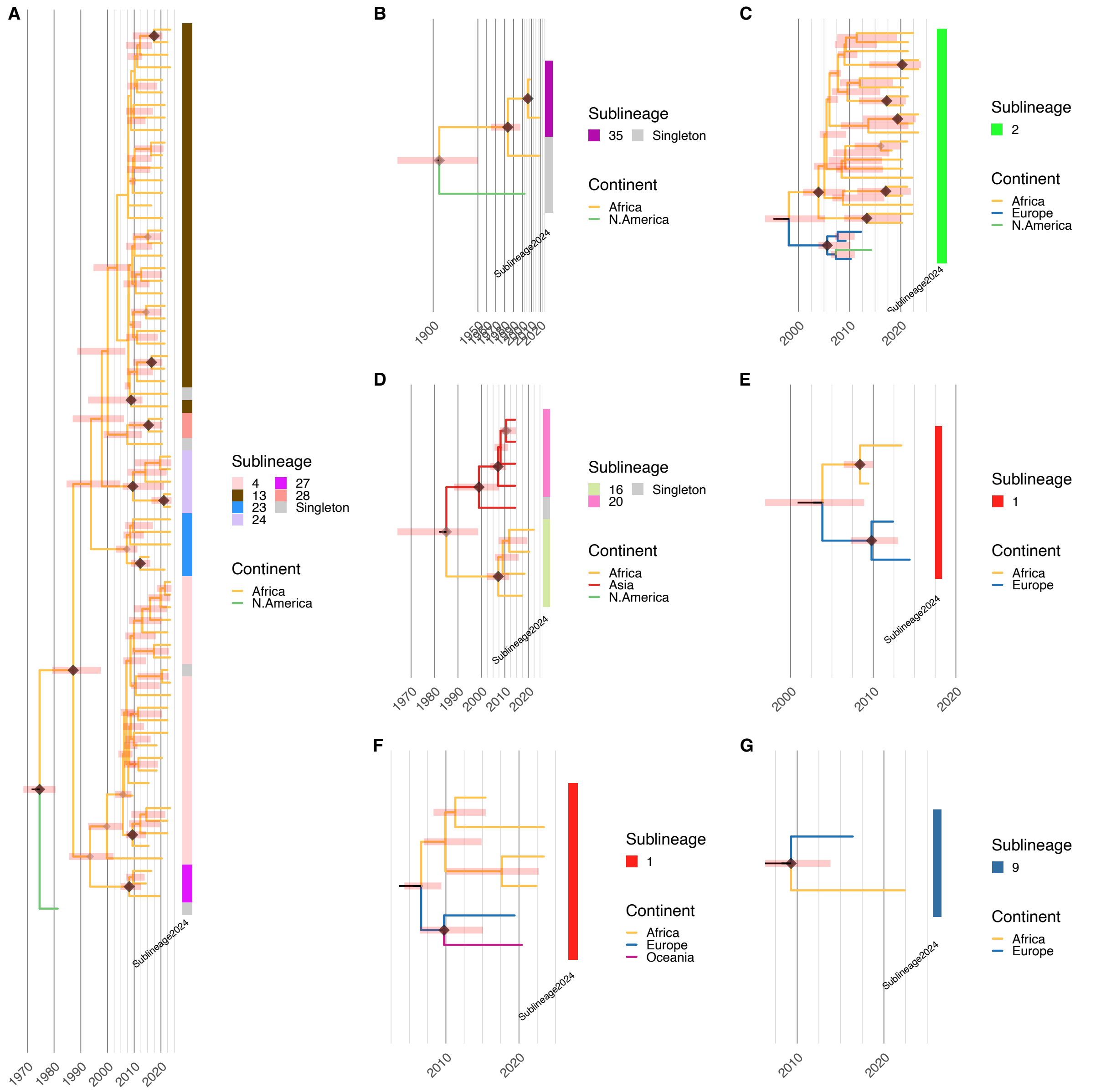


**Supplementary Figure 8. Temporal analysis of SS14 sublineages disseminating in Africa**. Time-scaled subtrees, showing date of most recent common ancestors (MRCAs) occurring in Africa, and sublineage assignments. Note, time-scaled trees were only possible for SS14-lineage.

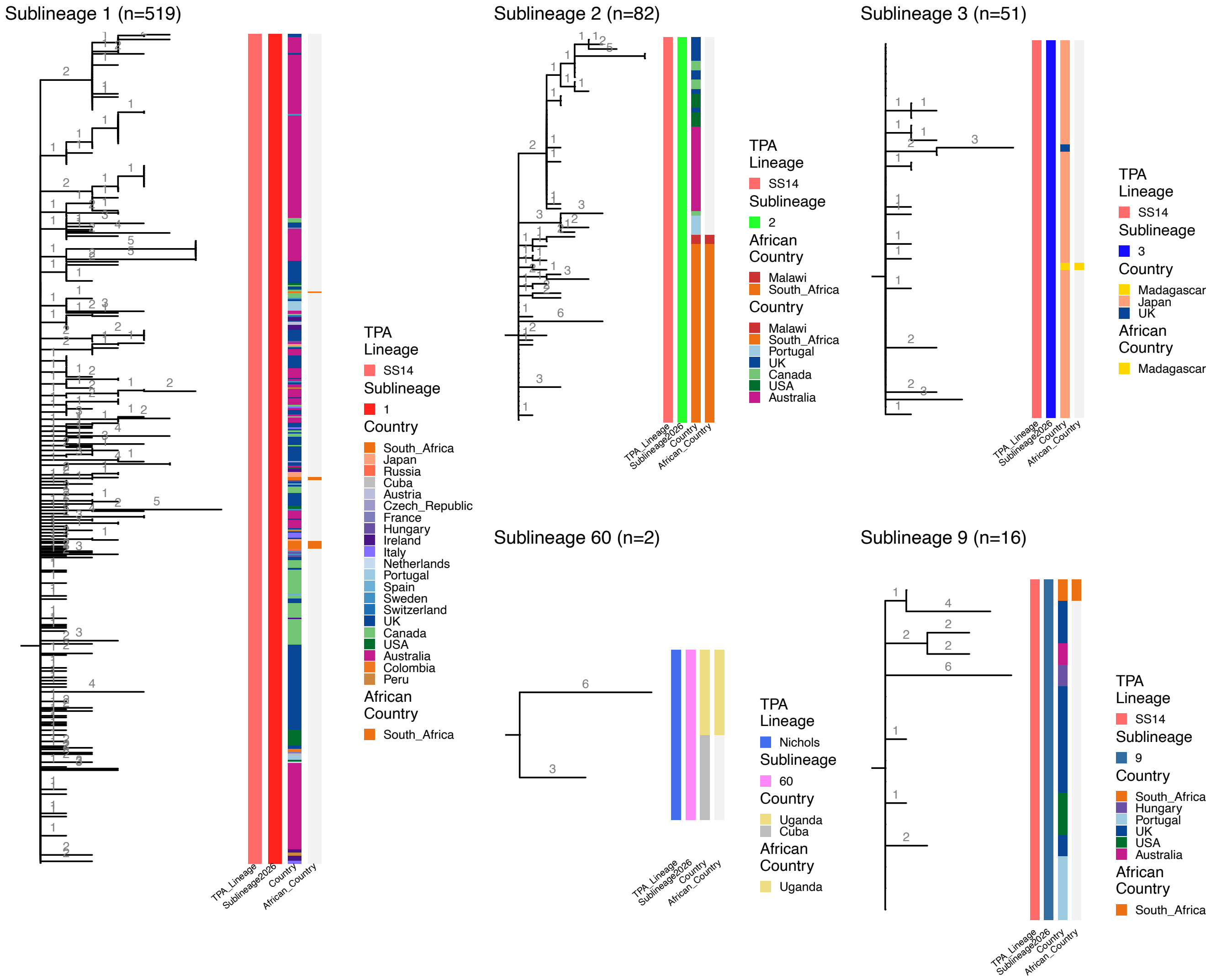


**Supplementary Figure 9.** Maximum likelihood subtree phylogenies for sublineages shared globally and found in Africa. Tree branches are scaled by number of substitutions (indicated by numbers). Coloured metadata tracks show Lineage, Sublineage, Country, and African Country.

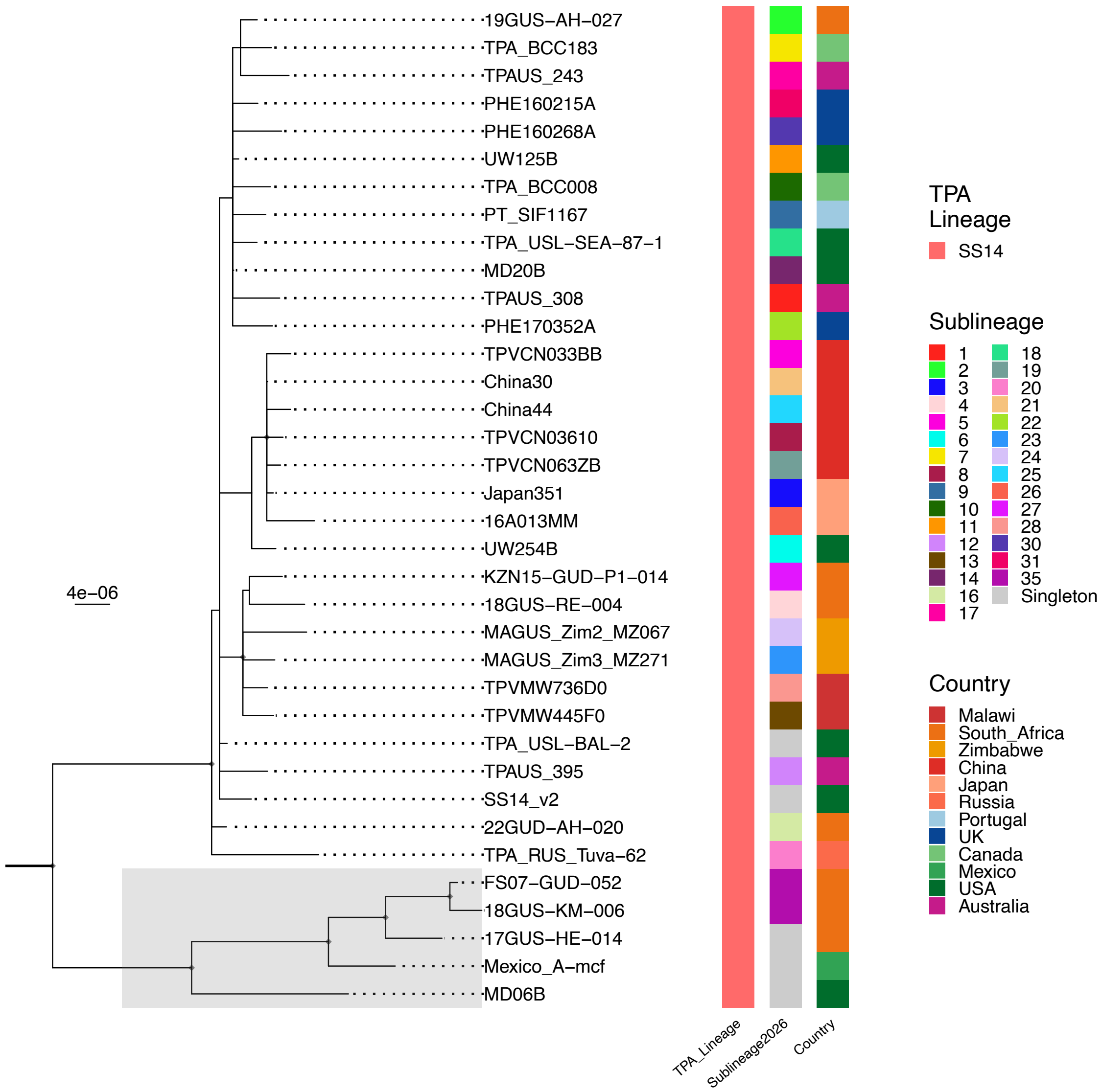


**Supplementary Figure 10. SS14 subtree highlighting Mexico_A-like genomes.** Maximum likelihood phylogenetic subtree (scaled in substitutions/site) comprising one randomly selected genome per sublineage and including all examples of the Mexico_A-like clade (shaded in grey). Coloured metadata tracks show Lineage, Sublineage and Country.

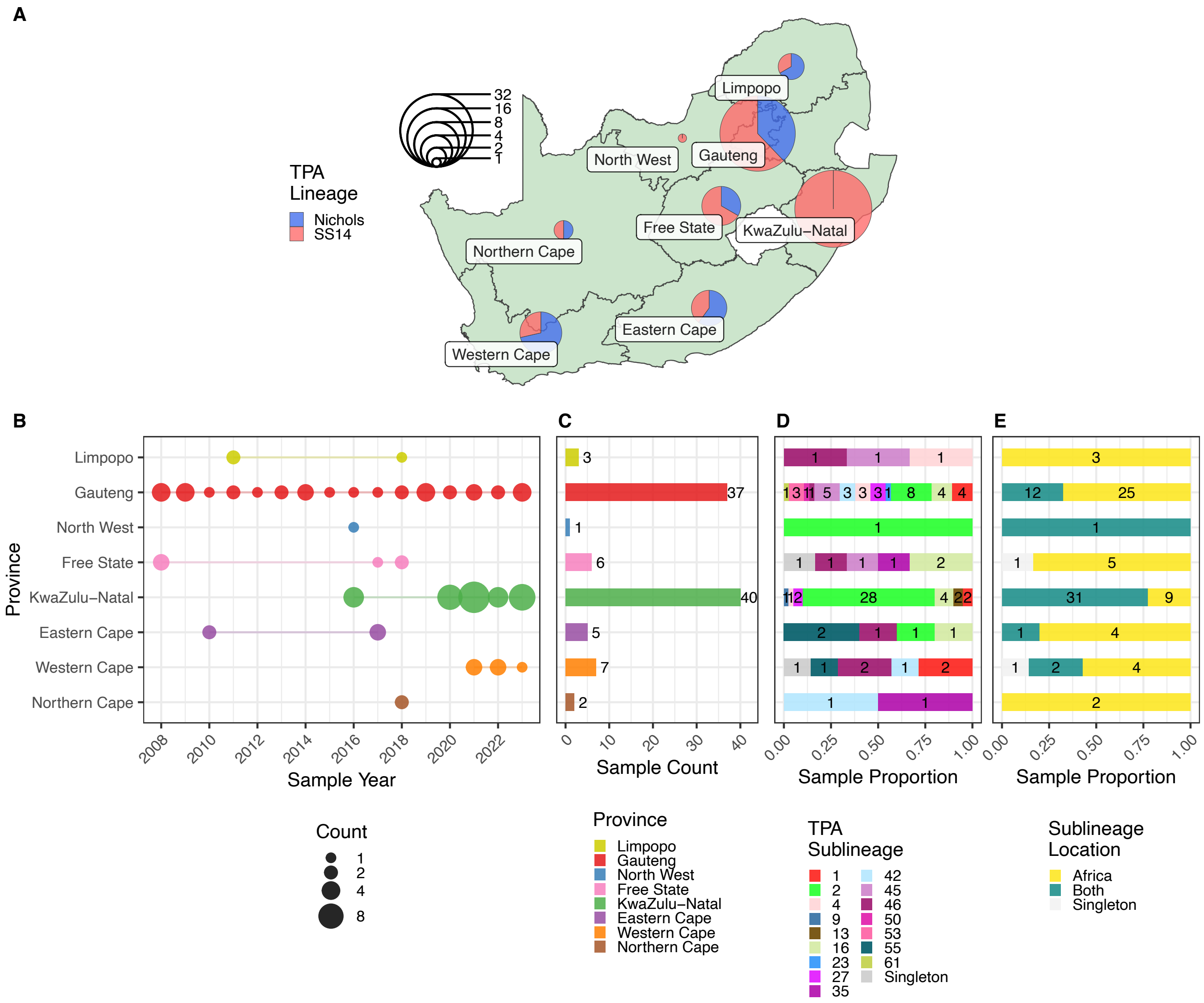


**Supplementary Figure 11.** A – Spatial distribution of TPA lineages in South Africa. B – Collection year of South African genomes by Province. C – Sample count per Province. D – Proportion of samples by sublineage. E – Proportion of samples by sublineage localisation (Africa, Africa+Global).

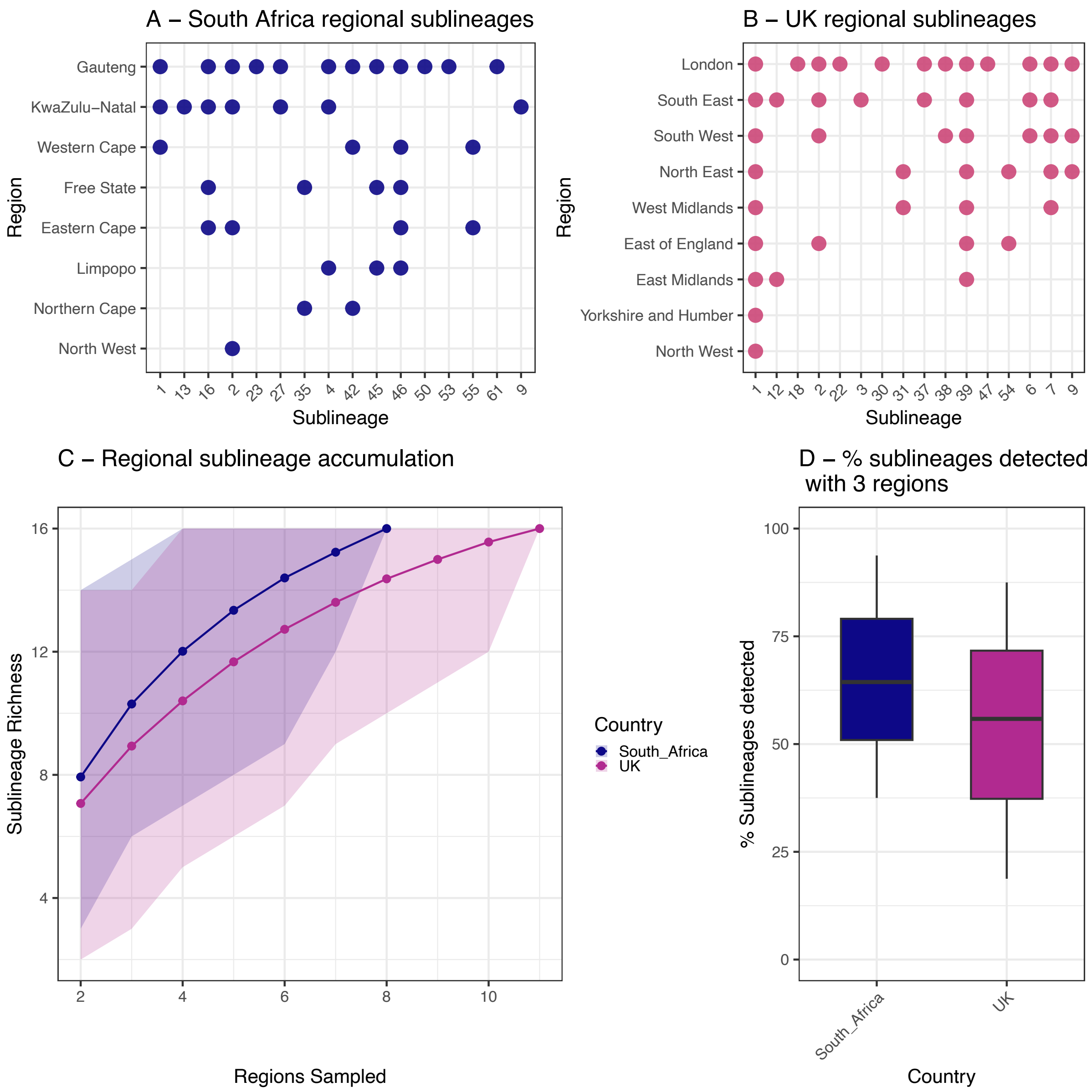


**Supplementary Figure 12.** Sampling three regions within a country captures 50% or more of sublineages, but broader sampling identifies further sublineages. A – Presence of sublineages by South African region. B – Presence of sublineages by UK region. C - Richness analysis, showing accumulation of sublineages within widely sampled countries (10,000 permutations). D – Percentage of sublineages detected when sampling only 3 regions (10,000 permutations)

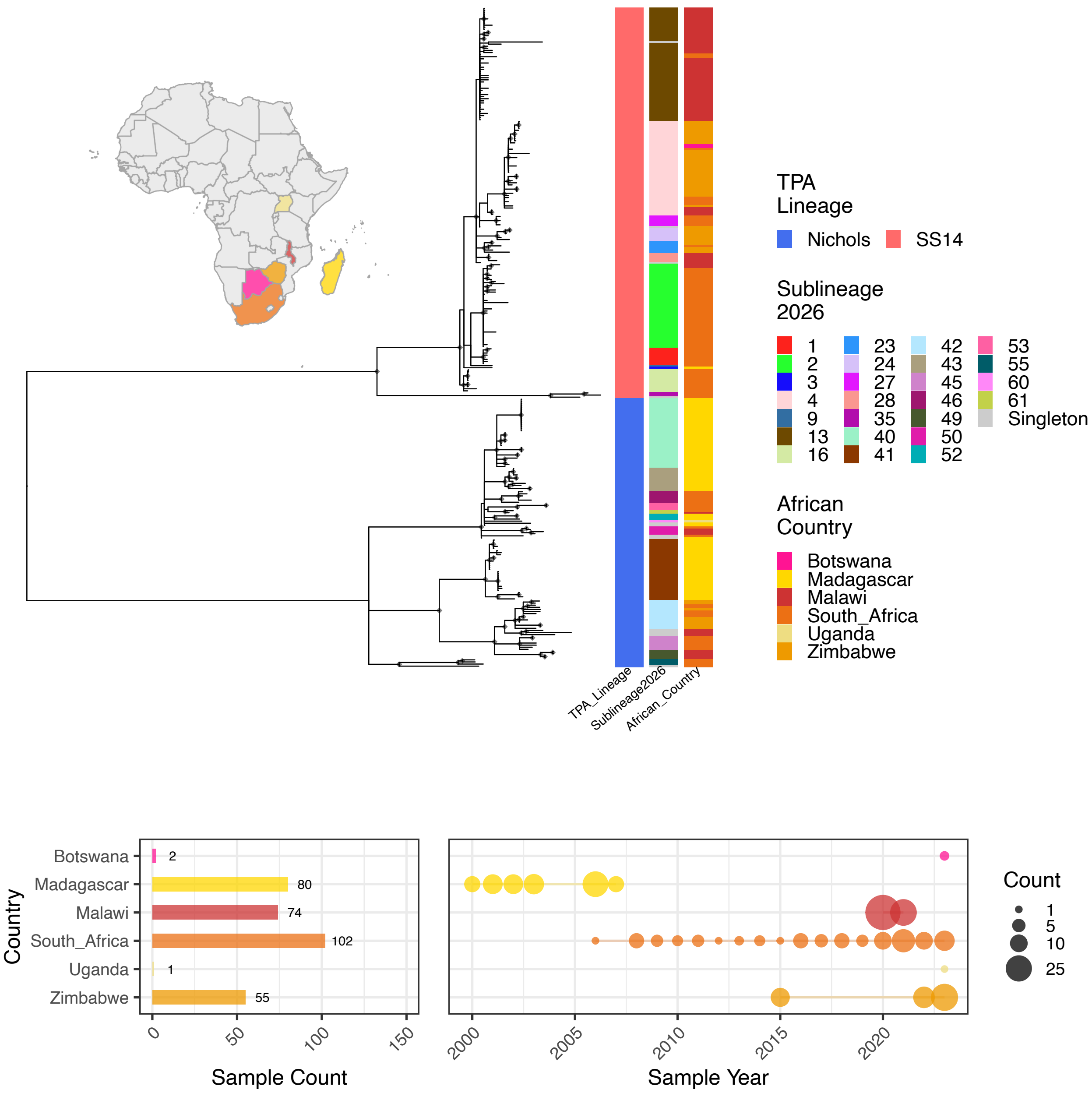


**Supplementary Figure 13.** A - Whole genome phylogeny of African *T. pallidum*, showing Lineage, Sublineage and Country. B – Sample count and year of sampling per country. Coloured tracks indicate TPA Lineage, Sublineage and Country.

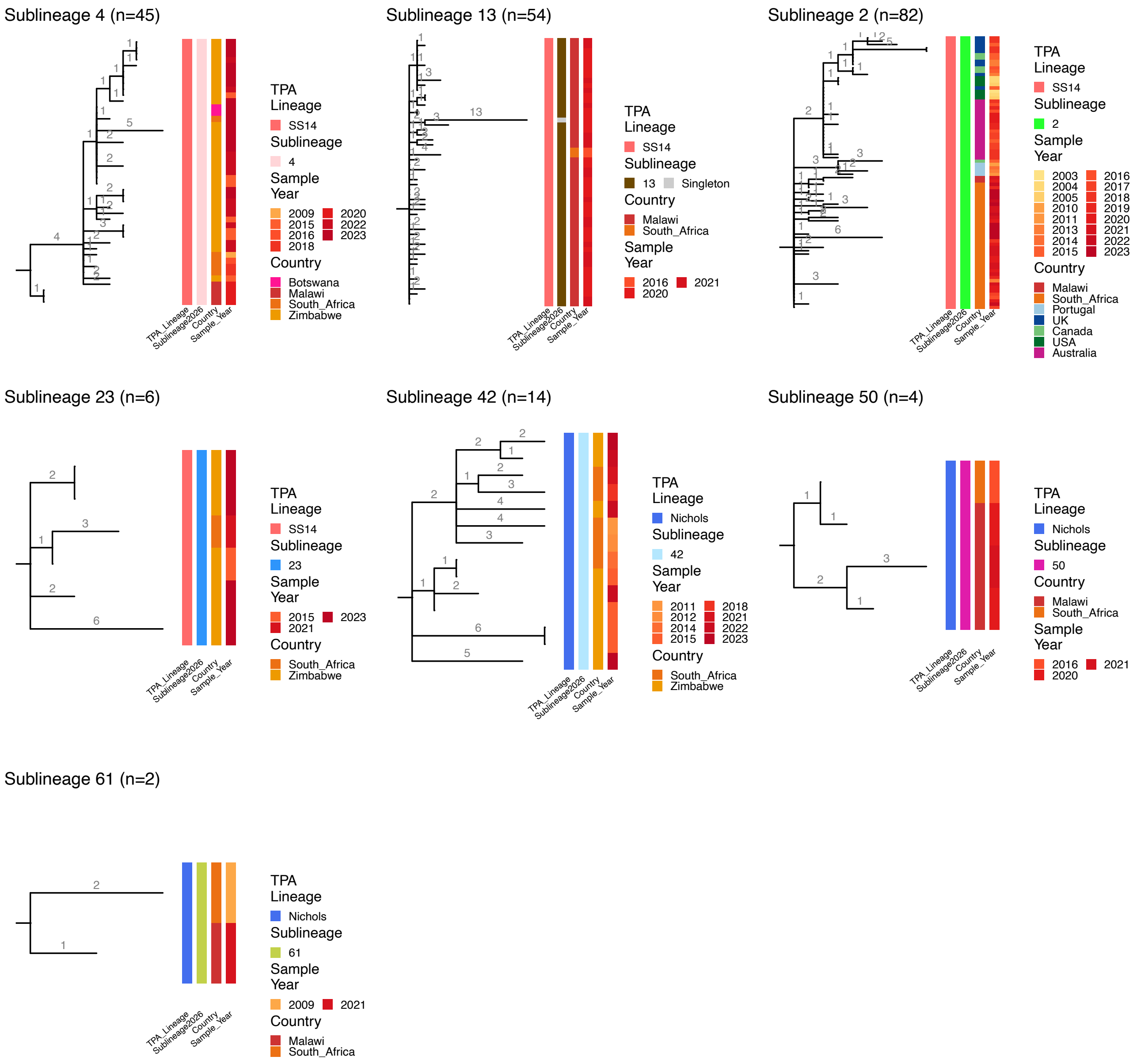


**Supplementary Figure 14.** Maximum likelihood subtree phylogenies for sublineages shared between multiple African countries. Tree branches are scaled by number of substitutions (indicated by numbers). Coloured metadata tracks show Lineage, Sublineage, Country, and Sample Year. Only one sublineage (2) was also found outside Africa.

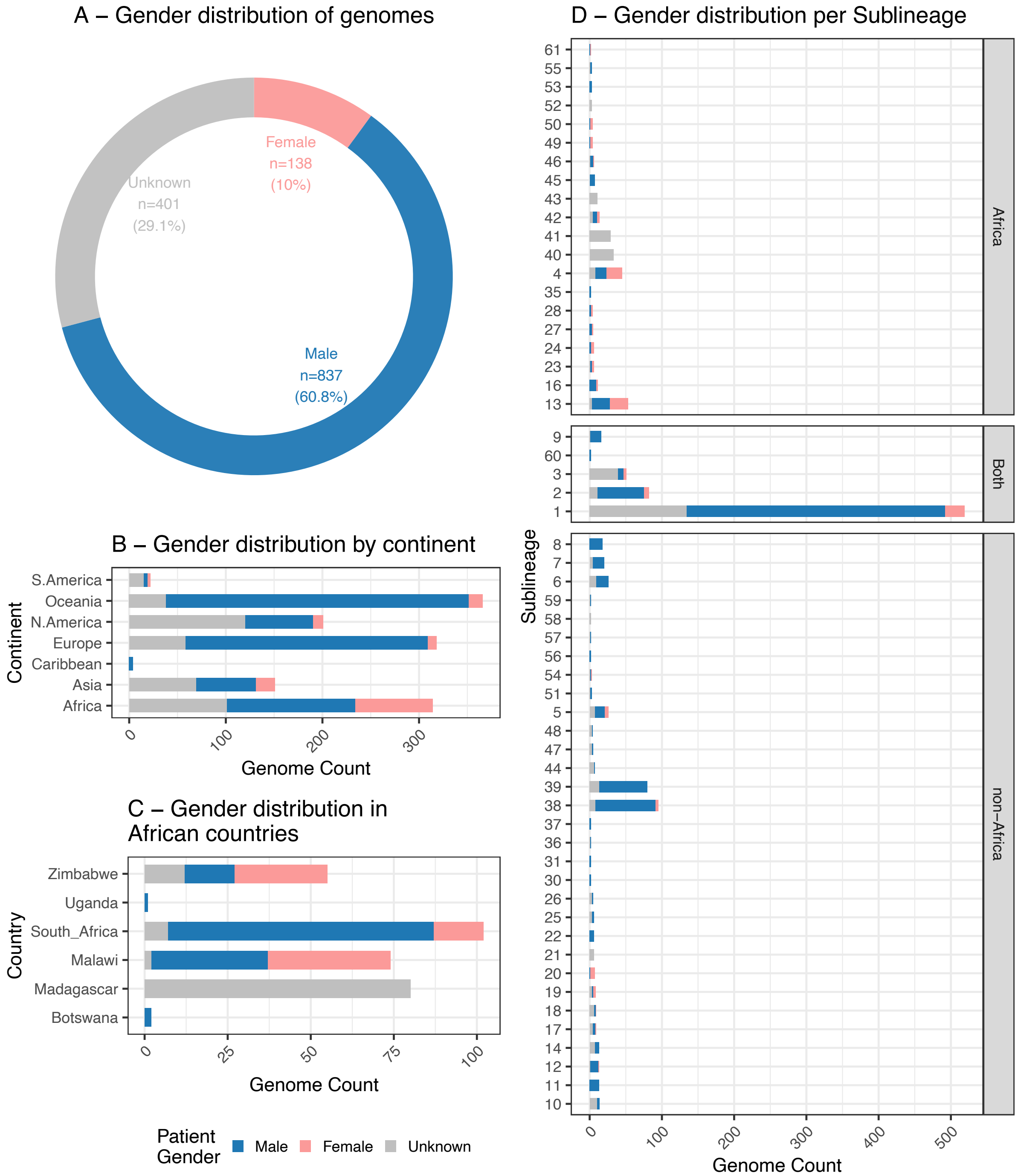


**Supplementary Figure 15.** Gender distribution amongst patients with whole genome sequences in this study. A – Overall gender distribution amongst full 1379 genomes. B – Gender distribution by Continent, C – Gender distribution amongst African countries. D – Gender distribution by TPA sublineage (facets indicate location sublineages detected).

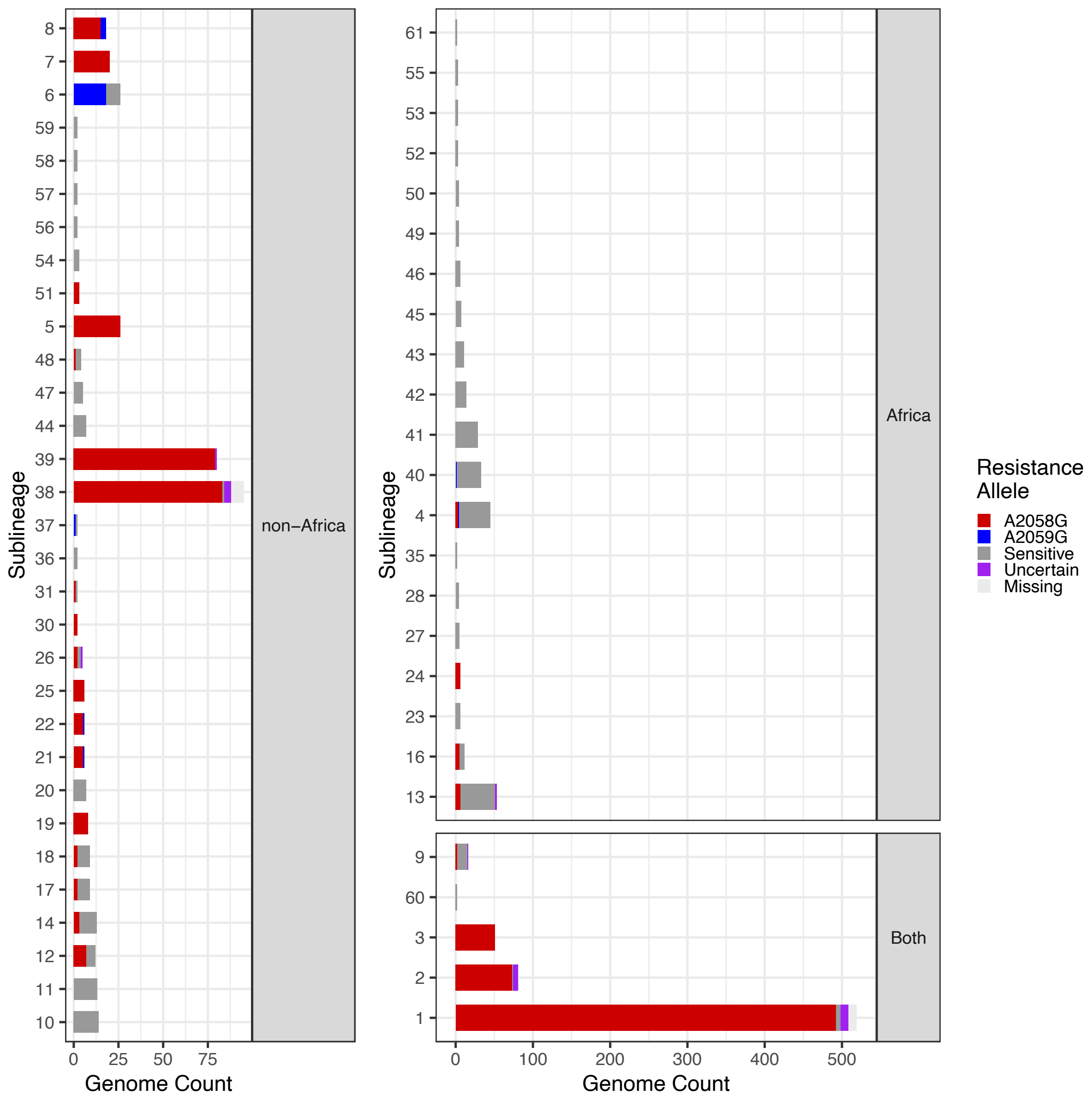


**Supplementary Figure 16.** Macrolide resistant sublineages are common globally but rare in Africa. Shows presence of macrolide resistance alleles (A2058G, A2059G) and wildtype by sublineage. Sublineages are grouped according to their region of detection (Africa, non-Africa, Both). Uncertain = mixed alleles at position. Missing = raw sequencing data unavailable for 23S locus.

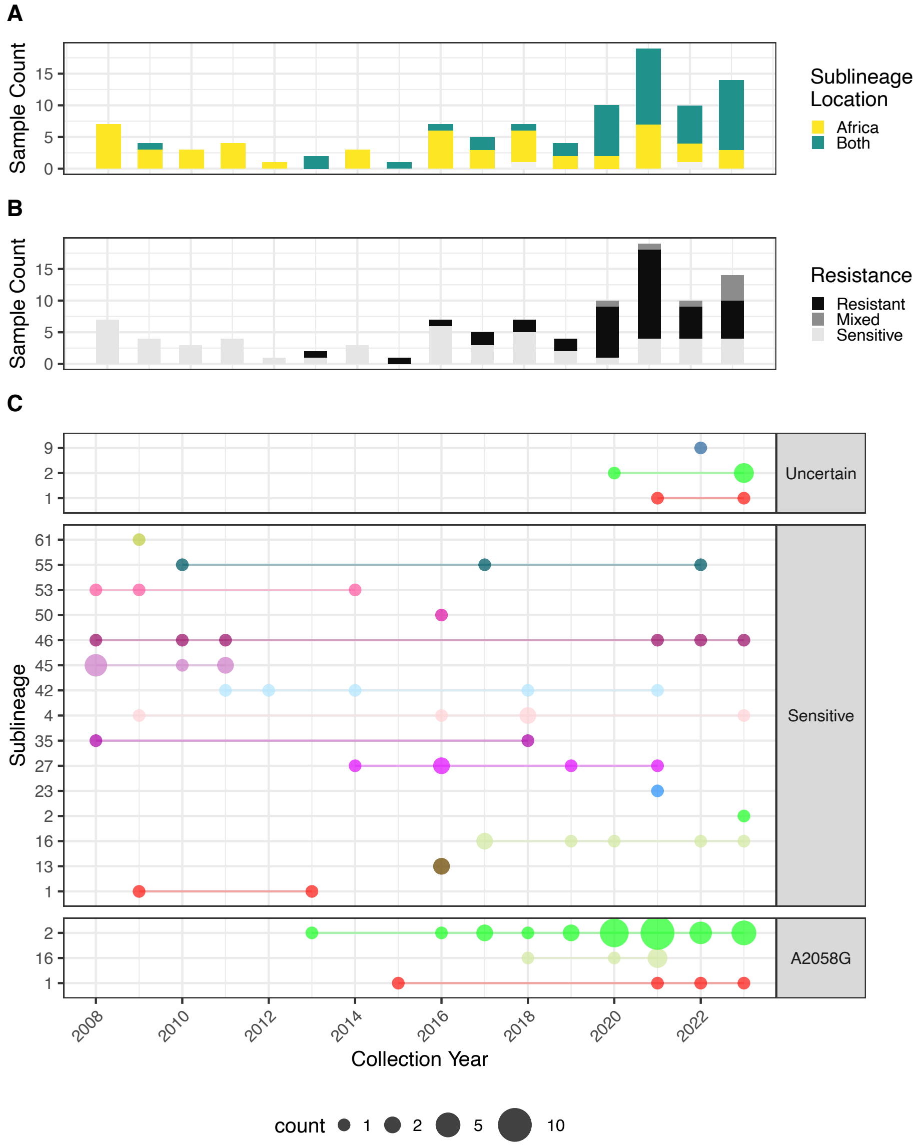


**Supplementary Figure 17.** Population dynamics of sublineages in South Africa show the emergence of macrolide resistant sublineages 1, 2 and 16 carrying A2058G. A – South African TPA genome count over time according to localisation of sublineage (African, African + Global). B – South African TPA genome count over time according to macrolide resistance allele. C – South African sublineages over time, according to whether genomes carry A2058G resistance allele.

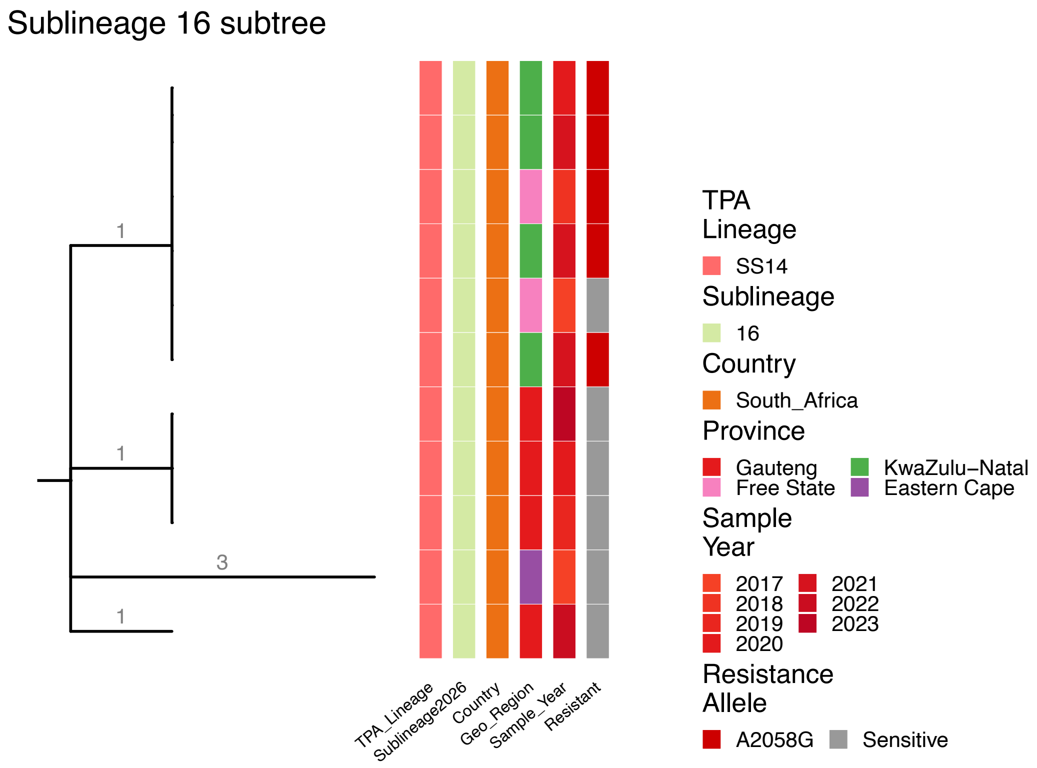


**Supplementary Figure 18.** Subtree of South Africa-specific sublineage 16. Coloured metadata tracks show Lineage, Sublineage, Country, South African Province, Sample Year, and macrolide resistance allele. Tree branches are scaled by number of substitutions (indicated by numbers).
